# Effects of the Histone Deacetylase Inhibitor Valproic Acid in Combination with Fear Memory Retrieval before Exposure Therapy for Spider Phobia: A Randomized Controlled Trial

**DOI:** 10.1101/2025.01.12.25320288

**Authors:** Nathalie S. Schicktanz, Frieder Dechent, Carlo Andreas Huber, Anja Zimmer, Galya Clara Iseli, Jeanne Howald, Maya Thalia Schenker, Johannes Gräff, Undine Lang, Dominique JF de Quervain, Dorothée Bentz

**Affiliations:** University of Basel, Division of Cognitive Neuroscience, Basel, Switzerland; University Psychiatric Clinics Basel, Basel, Switzerland; University of Basel, Research Cluster Molecular and Cognitive Neurosciences, Department of Biomedicine, Basel, Switzerland; École Polytechnique Fédérale Lausanne (EPFL), Laboratory of Neuroepigenetics, Brain Mind Institute, School of Life Sciences, Lausanne, Switzerland; École Polytechnique Fédérale Lausanne (EPFL), Synapsy Center for Neuroscience and Mental Health Research, School of Life Sciences, Lausanne, Switzerland

**Keywords:** Histone Deacetylase Inhibitor (HDACi), Fear Memory Retrieval, Remote Fear Memory Attenuation, Exposure Therapy, Phobic disorders

## Abstract

Return of fear after successful exposure therapy for phobia is a common clinical challenge. A previous study on mice demonstrated that the persistent attenuation of remote fear memories can be achieved by combining histone deacetylase inhibitors (HDACis) with fear-memory retrieval prior to extinction training. To evaluate the translational potential of this approach, we conducted a randomized, double-blind placebo-controlled trial. Forty-eight individuals with DSM-IV spider phobia received either HDACi valproic acid (VPA, 500 mg) or a placebo prior to the retrieval of fear memory followed by exposure therapy in virtual reality. No significant group difference was found in terms of behavioral change on the behavioral approach test at 3 months follow-up and baseline (primary outcome). However, the VPA group displayed significantly reduced fear in two self-report questionnaires related to spider phobia (Fear of Spider Questionnaire; Spider Phobia Beliefs Questionnaire) as compared to the placebo group. No group differences were observed for psychophysiological indicators of fear. The favorable impact of a single administration of VPA in combination with fear-memory retrieval prior to exposure therapy suggests that it might be an effective way to enhance symptom improvement at the subjective level in the treatment of phobias. Further studies need to investigate the conditions under which an improvement on the physiological and behavioral levels can be achieved as well.

## 1. Introduction

Specific phobias are a common mental disorder with an estimated lifetime prevalence of 12.5% (Michael et al., 2007). The symptomatology is characterized by excessive fear and avoidance behavior cued to specific objects or situations (American Psychiatric Association, 2013). Exposure therapy is the state-of-art treatment approach for specific phobias (Chambless and Ollendick, 2001). The hypothesized mode of action for fear reduction in exposure therapy is extinction (Hamlett et al., 2023), a process that establishes an alternative non-fear memory to compete with and suppress the original fear memory (Milad and Quirk, 2012). As a consequence, the original memory trace of the fear—although no longer expressed—remains intact and can return over time, upon context renewal, or when the original fearful object or situation is unexpectedly encountered (Bouton, 2004, 1993; Bouton and Bolles, 1979; Rescorla, 2004; Rescorla and Heth, 1975). The return of fear accordingly remains a common challenge in clinical practice, despite the success of fear reduction in short-term assessments following standard exposure therapy (Bentz et al., 2010; Craske and Mystkowski, 2006), so developing novel approaches to improve the stability of treatment results is of importance.

Targeting the learning and memory processes fundamental to exposure therapy represents a viable strategy. In this context, glucocorticoids with their memory-modulating properties have shown promising results in enhancing treatment success, potentially by weakening aversive- memory retrieval and strengthening extinction memories (Astill Wright et al., 2019; De Quervain et al., 2017, 2011). An alternative approach is to target the fear-memory reconsolidation process, especially if the goal is not only to enhance the efficacy of exposure therapy in reducing symptoms but also to increase the stability of treatment outcomes. Reconsolidation refers to the restabilization process of an already consolidated memory trace, which undergoes a brief phase of instability after retrieval (often referred to as memory reactivation). This process involves the synthesis of new proteins and RNA to stabilize the retrieved memory trace, presenting a window of opportunity to modify the existing memory trace permanently (Alberini, 2011; Przybyslawski and Sara, 1997). Whether the original or a modified memory trace is subsequently reconsolidated depends on various factors (for an overview, see Auber et al., 2013; Kindt and Elsey, 2023). These so-called boundary conditions include factors referring to the kind of retrieval cue and its timing but also to the fear memory itself and the approach used to target reconsolidation.

There are different approaches in use to destabilize and modify existing memories that can be divided into purely pharmacological or behavioral approaches or a combination of both. Pharmacological approaches involve administering substances that disrupt the reconsolidation process after retrieval (e.g., the protein synthesis inhibitor aniosomycin used in the seminal animal study by Nader et al. [2000] or propranolol in humans [Kindt et al., 2009]). Behavioral approaches refer to retrieval followed by the provision of new information. This approach was introduced by the retrieval-extinction paradigm in which extinction follows a short retrieval of a formerly acquired fear-memory trace (animals: Monfils et al., 2009; humans: Schiller et al., 2010). Both pharmacological and behavioral approaches have been found to successfully modify experimentally established fear memories in animals and humans using fear conditioning as the paradigm (Bentz and Schiller, 2015; Schwabe et al., 2014). The last decade has seen a large number of translational studies testing immediate symptom reduction based on either a behavioral or a pharmacological paradigm and the sustainability of these effects. For example, Soeter and Kindt (2015) describe how the application of propranolol after fear-memory retrieval in patients with spider phobia resulted in a greater symptom improvement compared to a placebo that persisted for at least one year after the procedure. However, the benefits of the propranolol reconsolidation paradigm have not been equally observed in all translational studies (Elsey et al., 2020; Steenen et al., 2016). For the behavioral approach, the majority of studies have looked at the enhancement of symptom reduction through retrieval followed by extinction-based therapy compared to standard exposure therapy. For this approach too, the findings are not unequivocal, as some studies have found improvements that lasted for up to 6 months (e.g., Björkstrand et al., 2017; Maples-Keller et al., 2017), while others found no effects (Shiban et al., 2015).

Taken together, attempts to target reconsolidation behaviorally or pharmacologically have not been consistently effective in clinical populations, whose symptoms are based on long-lasting memory traces such as those in phobic disorders. In this context, a study in mice showed that 1-day-old fear memories differ from remote fear memories acquired in the same classical-conditioning paradigm. This difference is not apparent in short-term outcomes (freezing) but becomes evident in long-term outcomes at a 30-day follow-up. Notably, only the remote-memory group exhibited a return of fear (Gräff et al., 2014). This finding is highly relevant, as efforts have primarily focused on translating findings from recent memories in the realm of reconsolidation research into clinical applications. The aforementioned study showed that whereas the recall of recent memories induced a brief period of hippocampal neuroplasticity, partly mediated by the S-nitrosylation of HDAC2 and histone acetylation, the recall of remote memories did not induce such neuroplasticity. Crucially, the same study showed that administering an HDAC2-targeting histone deacetylase inhibitor (HDACi) during the reconsolidation process effectively and persistently attenuated remote memories in a retrieval-extinction protocol (Gräff et al., 2014). The application of the HDACi was found to epigenetically prime the expression of neuroplasticity-related genes, enabling long-term modifications of remote memories.

Consequently, the application of HDACis in combination with the retrieval of a fear memory prior to extinction-based therapy may offer a novel therapeutic strategy for the persistent reduction of remote aversive memories. In the present study, valproic acid (VPA), a known HDACi, was combined with the retrieval of remote fear memories in individuals with spider phobia undergoing exposure therapy in virtual reality (VR). The purpose of this study was to evaluate whether VPA in combination with fear-memory retrieval enhances exposure therapy for phobias.

## 2. Experimental procedures

### 2.1. Study design and participants

We performed a double-blind, parallel-group (intervention vs. control), randomized controlled trial comparing the effect of the combination of VPA + retrieval with the effect of a placebo + retrieval prior to exposure therapy for a specific phobia (animal type: spiders). Healthy participants aged 18–40 years and diagnosed with a DSM-IV specific phobia (animal type: spiders) were recruited. To be eligible for the study, participants were required to have a score of 1–7 points on a real-life behavioral approach test (BAT in vivo) to ensure their willingness to subject themselves to exposure and to prevent ceiling effects post intervention. The study protocol of the clinical trial (clinicaltrials.gov, identifier: NCT02789813), including the definition of the primary and secondary outcomes and the plan for the statistical analysis was approved by the Ethics Committee of Northwest and Central Switzerland (EKNZ) before the start of the study. The entire study was performed in accordance with the Declaration of Helsinki. All participants gave written informed consent to trial participation. Participants received a compensation of CHF 250 for participating in the trial.

### 2.2. Randomization and masking

Participants were randomly assigned by the experimenter to treatment groups (VPA + retrieval or placebo + retrieval), with stratification by sex. The randomization lists and study medication were prepared by the hospital pharmacy at the University Hospital of Basel in Switzerland. Block randomization in groups of four was used. Each block included two allocations for each treatment condition. Visually identical syrups containing VPA or placebo filled in 20 ml glass bottles with identical labels were provided. These measures ensured the concealment of assignments from the enrolling physician, experimenter, and participants, maintaining a double-blind approach regarding VPA administration. Additionally, the personnel processing psychological and physiological data remained unaware of participant group assignments.

### 2.3. Study tasks

#### 2.3.1. Retrieval

Participants were shown a virtual spider for 5 seconds in a virtual room (see Figure 2) to initiate retrieval of the fear memory. This was done 10 minutes prior to exposure therapy to target the reconsolidation window (Schiller et al., 2010). Afterward, participants were asked to indicate the fear they experienced via a Subjective Units of Distress Scale (SUDS) (0 = no fear, 10 = maximum fear) (Wolpe, 2018). Electrodermal activity (EDA) and electrocardiography (ECG) were measured during the 5 seconds of the presentation of the virtual spider.

**Figure 1.**
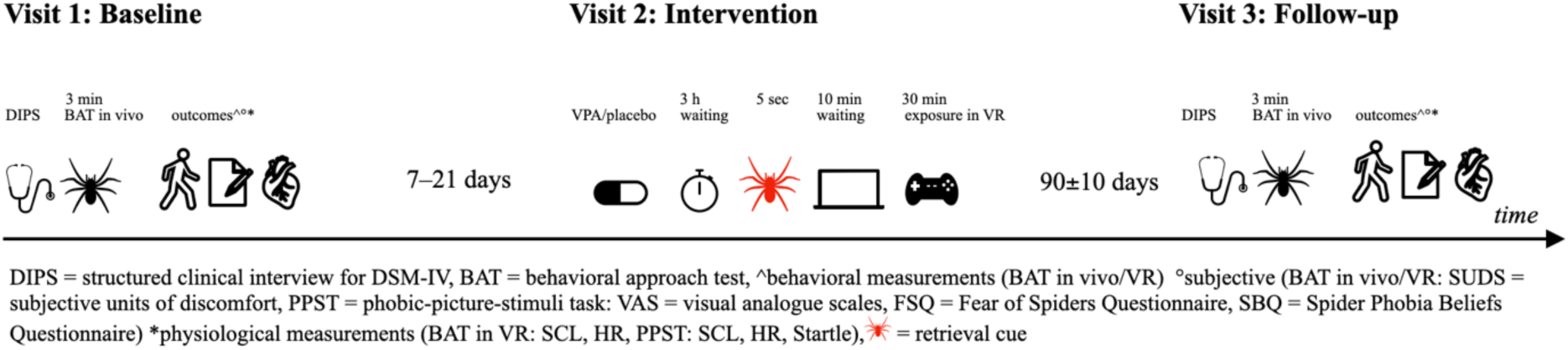
Study schedule. This figure illustrates the sequence of the study tasks for obtaining the primary and secondary outcome measurements at baseline (visit 1) and follow-up (visit 3), as well as the sequence of the intervention during visit 2.

**Figure 2.**
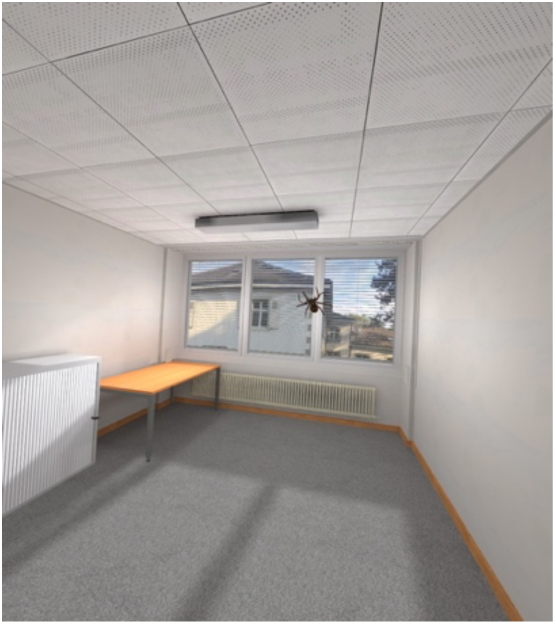
Virtual environment (VR). The picture is a screen shot taken from scene 1 and 10 of the VR exposure paradigm. It also corresponds to what the participants saw for the 5 seconds that the retrieval cue was presented in VR.

#### 2.3.2. Exposure in VR

For exposure therapy, we used the 30-minute hierarchical VR-exposure protocol employed in the study by Shiban et al. (2015). During each of the ten 3-minute scenes in VR, participants rated their fear on the SUDS at the beginning and end of each scene. Minimal verbal support was given by the experimenter, who also checked for simulation sickness and recorded the SUDS measurements. Psychophysiological measurements (EDA, ECG) were continuously taken during exposure.

#### 2.3.3. BAT in vivo

We used a BAT to measure approach behavior to a living house spider (*Tegenaria atrica*, size: approx. 5 cm). This BAT in vivo corresponded to the one used by Lass-Hennemann and Michael (2014). Before starting the BAT, participants rated their expected fear during the BAT using a SUDS (0 = no fear, 10 = maximum fear imaginable). The BAT assessed participants’ willingness to approach a spider in a sealed transparent container following 12 graded steps (cumulative points per step in brackets): refuses to enter the room (0), stops at 5 m (1), 4 m (2), 3 m (3), 2 m (4), 1 m (5), near the container (6), touches the container (7), removes the lid (8), puts a hand inside (9), touches the spider (10), holds the spider (<20s = 11, ≥20s = 12). The task ended when the participant could not proceed, completed the last step, or after 3 minutes. At the final position reached, participants rated their actual fear with a SUDS.

#### 2.3.4. BAT in VR

We used a BAT to measure approach behavior to a VR spider. Participants had 3 minutes to approach the virtual spider. Before these 3 minutes began, their position was fixed with the virtual spider in their field of vision for 15 seconds to record SUDS and physiological fear indicators (EDA, ECG) in a standardized way. Approach behavior was determined by means of virtual distance to the spider at the end of the BAT in VR.

#### 2.3.5. Phobic-picture-stimuli task (PPST)

The phobic-picture-stimuli task (PPST) was designed to obtain subjective and psychophysiological reactions to fear-related (spider), negative (snake), and neutral stimuli. Half of the presented pictures in each category were combined with startle probes (white noise, 100 dB). The duration of the main phase of the PPST was approximately 20 minutes.

### 2.4. Outcomes

Our primary outcome was change in approach behavior as measured by the difference in the BAT in vivo scores between visit 1 (baseline) and visit 3 (follow-up) (BAT difference score). The scores ranged from 0 to 12 (higher scores indicate higher approach behavior).

Our secondary outcomes were the differences between visit 1 (baseline) and visit of 3 (follow-up) (difference score) in: (1) the SUDS after BAT in vivo, (2) the BAT in VR, (3) the SUDS after BAT in VR, (4) heart rate (HR) after BAT in VR, (5) skin conductance levels (SCL) after BAT in VR, (6) the valence ratings of spider pictures, (7) the arousal ratings of spider pictures, (8) the anxiety ratings of spider pictures, (9) HR while looking at spider pictures, (10) SCL while looking at spider pictures, (11) startle reactions to spider pictures, (12) the Fear of Spider Questionnaire (FSQ; sum score ranges from 0 to 108) (Szymanski and O’Donohue, 1995), (13) the Spider Phobia Beliefs Questionnaire (SBQ; mean sum score 0–100) (Arntz et al., 1993), (14) State–Trait Anxiety Inventory state version (STAI-S) (Spielberger, C.D. et al., 1970), (15) DSM-IV impairment scores, and (16) DSM-IV distress scores (for details on the secondary outcomes, see the supplementary information).

### 2.5. Statistical analyses

We conducted a per-protocol analysis using R Studio version 4.2.3 (R Core Team, 2021). Linear models were applied to assess the impact of our interventions. These models incorporated ANOVA (type II sums of squares) and were used with dependent variables representing the differences in the measurements at visit 3 and visit 1. The covariates included age and sex, while group allocation served as an independent variable. Effect sizes were calculated using Cohen’s *d*. The model assumptions were checked using the Performance package in R (Lüdecke et al., 2021) and confirmed for the main analysis (for additional analysis, see the supplementary information). The significance threshold was set at *p* < 0.05 for the primary outcomes and *p* < 0.003125 (Bonferroni correction for 16 independent tests) for the secondary outcomes. We aimed to detect large drug effects (*d* = 0.8) with 80% power and α = 0.05. A power analysis (G*Power 3.1) indicated 25 participants per group was the minimum group size to measure statistically relevant effects.

### 2.6. Deviations from initial protocol

The approved study protocol foresaw four experimental groups with a total of 100 participants: intervention (VPA + retrieval), control I (placebo + retrieval), control II (VPA + no retrieval), and control III (placebo + no retrieval). To ensure that we would be able answer our main question of the impact of VPA + retrieval on exposure therapy for phobias even in case of recruitment difficulties, we first recruited participants for the intervention and control I. Due to slow recruitment, we conducted an interim analysis with 48 participants. The study was terminated after the interim analysis on May 24, 2018, as it did not support our primary hypothesis that VPA + retrieval would lead to greater approach behavior as measured in the BAT in vivo compared to placebo + retrieval. The data from the participants recruited for control II and III were not analyzed and are not shown in the flowchart, see Figure 3.

**Figure 3.**
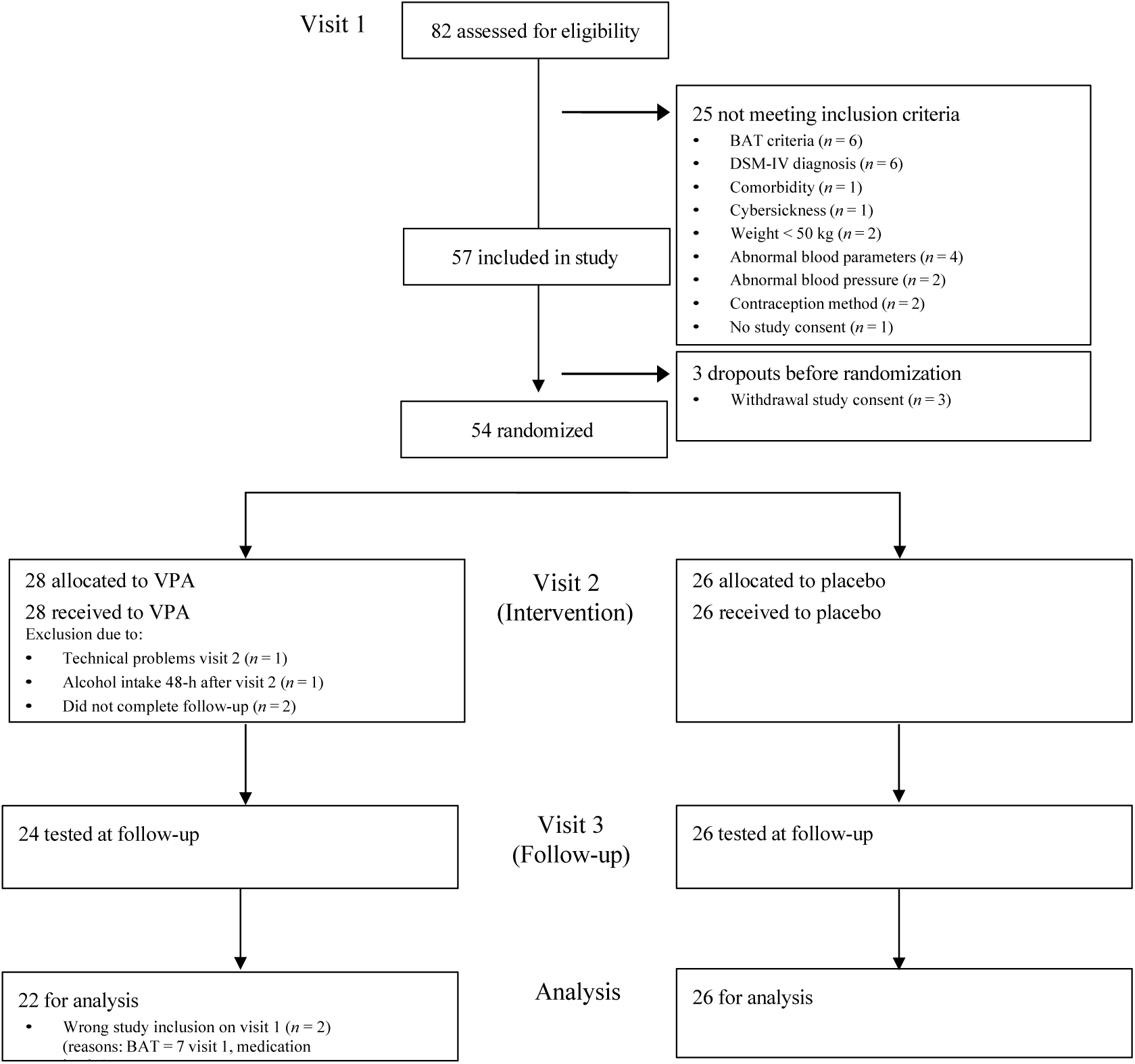
Flowchart of participants: CONSORT. The flowchart depicts the participants’ progression through the study from inclusion (visit 1) to data analysis for the participants recruited for the VPA + retrieval and placebo + retrieval groups. BAT = behavioral approach test, VPA = valproic acid.

## 3. Results

### 3.1. Participants’ characteristics and study flow

Eighty-two individuals were screened for trial participation. Fifty-four participants were randomized (28 received VPA, 26 received a placebo), 50 participants were tested at visit 3, and 48 participants were included in the analysis. For full information about the participants’ study flow, see Figure 3; for the participants’ characteristics, see Table 1.

**Table 1.** Participants’ characteristics.

| **Outcome** | **Placebo** | **VPA** |
| --- | --- | --- |
| Sex (female / male) | 23 / 3 | 20 / 2 |
| Comorbid specific phobia (y / n) | 11 / 15 | 8 / 14 |
| DSM-IV diagnoses (animal type: spider) at follow-up (y / n) | 19 / 7 | 14 / 8 |
| Age (*SD*) | 24.42 (4.44) | 24.41 (4.33) |
| Body mass index (*SD*) | 24.05 (3.88) | 23.31 (3.18) |

### 3.2. Retrieval

There was no significant group difference in the SUDS ratings after the retrieval procedure (*p* = 0.11, placebo: mean = 4.38, *SD* = 2.64, range 0–9; VPA: mean = 5.54, *SD* = 2.06, range 2–9). Furthermore, there was no significant group difference in the EDA and ECG measurements (all *p* > 0.37).

### 3.3. Effect of VPA on primary and secondary outcomes

Group allocation did not have a statistically significant impact on the primary outcome, the difference in approach behavior as measured by the BAT in vivo (*F* (1, 44) = 0.56, *p* = 0.46, *d* = 0.23). For the secondary outcomes, Bonferroni-corrected significant differences were observed between the groups in two of the 16 secondary outcome measures, both of which were self-report questionnaires assessing the fear of spiders: FSQ (*F* (1, 44) = 9.78, *p* = 0.003, *d* = −0.95), and the SBQ (*F* (1, 43) = 11.09, *p* = 0.002, *d* = −1.02) (see Figure 4 and Table 2). These findings indicate a more pronounced reduction in the subjective experience of phobic fear in the VPA + retrieval group compared to the placebo + retrieval group. None of the other secondary outcomes showed a significant group-allocation difference after a Bonferroni correction (*p* ≥ 0.019; see the supplementary information).

**Figure 4.**
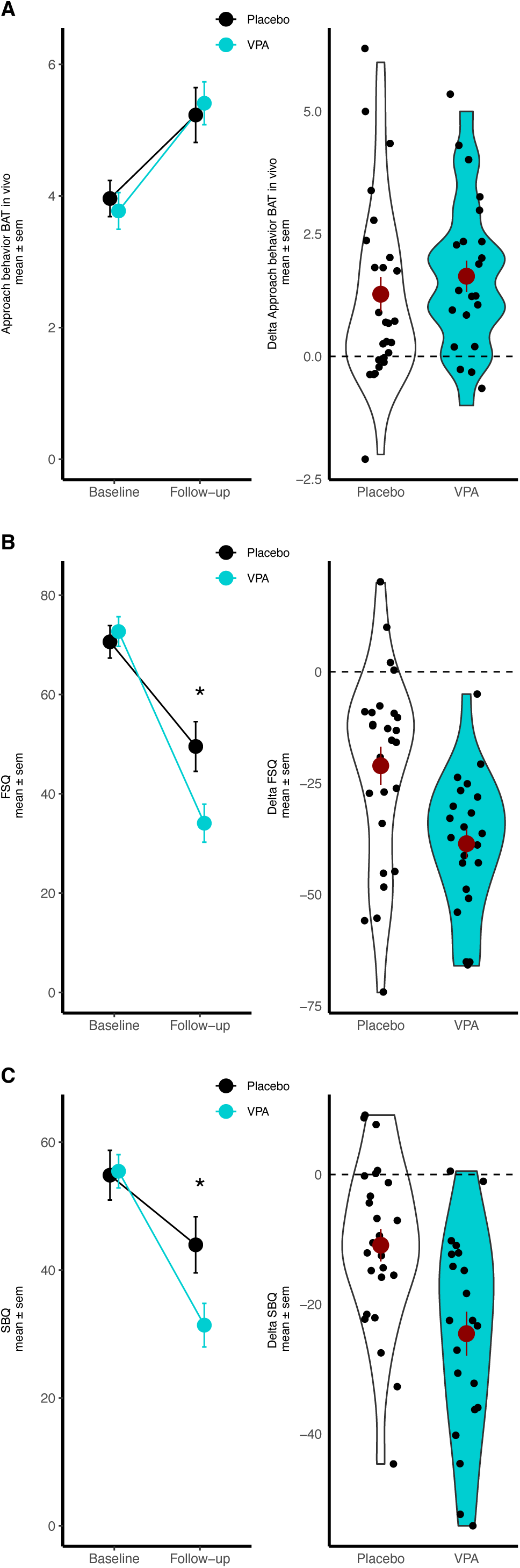
Primary and significant secondary outcomes at visit 1 (baseline) and visit 3 (follow-up) The black line represents the placebo + retrieval group, and the turquoise line represents the valproic acid + retrieval group. Displayed are the means and standard errors of the means. BAT in vivo = behavioral approach test in vivo (0–12), FSQ = Fear of Spiders Questionnaire (sum score ranges from 0 to 108), SBQ = Spider Phobia Beliefs Questionnaire (mean sum score ranges from 0 to 100). On the left side, the raw values of both visits are displayed. On the right side, the delta outcome variables between follow-up and baseline are displayed.

**Table 2.** Descriptive statistics of primary and significant secondary outcomes, effect sizes, and *p* values.

| Outcome | Time | Group | *n* | Mean | Median | *SD* | Min | Max | Cohen's *d* | *p* | Effect size |
| --- | --- | --- | --- | --- | --- | --- | --- | --- | --- | --- | --- |
| Approach behavior BAT in vivo | Baseline | Placebo | 26 | 3.96 | 4.00 | 1.40 | 1.00 | 6.00 |  |  |  |
|  | Baseline | VPA | 22 | 3.77 | 3.50 | 1.31 | 1.00 | 6.00 | −0.14 |  |  |
|  | Follow-up | Placebo | 26 | 5.23 | 5.00 | 2.12 | 1.00 | 8.00 |  |  |  |
|  | Follow-up | VPA | 22 | 5.41 | 5.00 | 1.53 | 2.00 | 8.00 | 0.10 |  |  |
|  | Difference score | Placebo | 26 | 1.27 | 1.00 | 1.80 | −2.00 | 6.00 |  |  |  |
|  | Difference score | VPA | 22 | 1.64 | 1.50 | 1.50 | −1.00 | 5.00 | 0.22 | 0.46 | *d* = 0.23 |
| FSQ | Baseline | Placebo | 26 | 70.62 | 70.50 | 16.74 | 37.00 | 97.00 |  |  |  |
|  | Baseline | VPA | 22 | 72.68 | 71.00 | 14.08 | 45.00 | 99.00 | 0.13 |  |  |
|  | Follow-up | Placebo | 26 | 49.54 | 50.00 | 25.53 | 3.00 | 86.00 |  |  |  |
|  | Follow-up | VPA | 22 | 34.09 | 30.00 | 17.97 | 6.00 | 75.00 | −0.69 |  |  |
|  | Difference score | Placebo | 26 | −21.08 | −14.00 | 21.74 | −72.00 | 20.00 |  |  |  |
|  | Difference score | VPA | 22 | −38.59 | −36.50 | 15.34 | −66.00 | −5.00 | 0.92 | 0.003 | *d* = 0.95 |
| SBQ | Baseline | Placebo | 26 | 54.84 | 59.58 | 19.85 | 4.17 | 90.54 |  |  |  |
|  | Baseline | VPA | 21<sup>a</sup> | 55.45 | 51.35 | 11.96 | 31.04 | 77.71 | 0.04 |  |  |
|  | Follow-up | Placebo | 26 | 43.94 | 47.92 | 22.38 | 0.83 | 80.42 |  |  |  |
|  | Follow-up | VPA | 22 | 31.38 | 31.88 | 16.00 | 10.63 | 71.88 | −0.64 |  |  |
|  | Difference score | Placebo | 26 | −10.89 | −10.64 | 12.75 | −44.63 | 9.17 |  |  |  |
|  | Difference score | VPA | 21<sup>a</sup> | −24.53 | −22.50 | 15.49 | −54.17 | 0.52 | 0.97 | 0.002 | *d* = 1.02 |

VPA = valproic acid, FSQ = Fear of Spiders Questionnaire (sum score ranges from 0 to 108), SBQ = Spider Beliefs Questionnaire (mean sum score ranges from 0 to 100), *SD* = standard deviation, min = minimum, max = maximum, Cohen's *d* refers to effect size calculated based on mean values, *p* refers to *p* value of main effect of group allocation in the used statistical model, i.e., difference between follow-up and baseline. Effect size refers to effect size of main effect of group allocation in the used statistical model, i.e., statistical model including age and sex as covariates (for more information, see the supplementary information).
<sup>a</sup> SBQ was not collected for one participant at baseline.

### 3.4. Valproic acid in blood levels before and after study-medication intake at visit 2

Before study-medication intake, all the participants had undetectable levels of VPA in their blood (the laboratory sensitivity was ≥ 2.8 mg/l, values labeled by the laboratory with < 2.8 mg/l were set to 0). Following the intake of the study medication, the blood levels for the placebo + retrieval group remained at zero (mean = 0, *SD* = 0). For the VPA group, there was a notable increase in blood levels (mean = 40.27, *SD* = 6.28, min = 25.20, max = 49.60, unit = mg/l). This increase is significantly different from zero (one-sample *t* test against zero, *t* (19) = 28.69, *p* < 2.2E-16).

### 3.5. Participants’ perception of intake of VPA or placebo

There were no significant group differences in the participants’ perception of having received VPA or placebo when asked at visit 2 or visit 3 (*p* ≥ 0.18) (for more information, see the supplementary information).

### 3.6. Adverse events

There were no significant group differences in the occurrence of adverse events at visit 2 after medication intake (*p* > 0.49). Eleven participants from the VPA + retrieval group reported 15 adverse events overall, while 14 participants from the placebo + retrieval group reported 23 adverse events at visit 2 after medication intake (see Supplementary Table ST4 and Supplementary Table ST5).

## 4. Discussion

The present study found no significant differences in the primary outcome—approach behavior as measured in a real-life BAT conducted 3 months after the intervention—between the group that received VPA in combination with retrieval prior to exposure therapy versus the group that received a placebo in combination with retrieval prior to exposure therapy. However, noteworthy findings were seen in the secondary outcomes, where two out of the 16 outcomes displayed significant effects after a Bonferroni correction. Specifically, these two secondary outcomes revealed a significant reduction in subjective fear of spiders as assessed by the Fear of Spiders Questionnaire (FSQ) and the Spider Phobia Beliefs Questionnaire (SBQ) in the group that received VPA in combination with retrieval as compared to the group that received a placebo in combination with retrieval. This reduction was observed in addition to the effects of exposure therapy itself, which exhibited a medium effect size in the control group (generalized semipartial R2, FSQ: 0.22, SBQ: 0.11). The FSQ and SBQ were used to assess self-reported emotions and anticipated reactions in the presence of spiders, including cognitive aspects of these emotions. Importantly, there were no significant differences in the participants’ perceptions of whether they had received VPA or a placebo. This indicates that the subjective emotional and cognitive responses were not biased by the participants’ perceptions of their treatment allocation.

Beyond the VPA effects on fear as assessed with the two questionnaires, no additional effects were observed in the participants’ actual fear responses in the presence of a living spider or a virtual one, in their psychophysiological reactions, in DSM-IV impairment and distress criteria, or in their general state anxiety. Since our primary and secondary outcomes assessed distinct aspects of phobic fear, our results may indicate that the proposed mechanism of action of the study medication did not affect all of these aspects similarly following a single dose of medication. LeDoux and Pine (2016) argue that responses to threats should be differentiated into behavioral and subjective components. They propose that two distinct brain circuits underlie (1) behavioral responses, which are accompanied by physiological changes in the brain and body, and (2) conscious feeling states, as reflected in self-reports of fear and anxiety. It is possible that the outcomes from the subjective self-report fear questionnaires were more sensitive to the treatment compared to behavioral components. In line with this explanation, a single exposure therapy session showed an effect on self-report questionnaires but not on approach behavior as measured in a BAT in vivo (Bentz et al., 2021; Zimmer et al., 2021). It thus remains to be investigated whether different dosages or repeated administrations of VPA in combination with exposure therapy might also affect objective measures in addition to the subjective measures.

During the exposure sessions, the VPA and placebo groups did not exhibit significant differences in terms of SUDS, SCL, and HR at the beginning and end of each exposure scene level and when comparing the beginning and end of the complete exposure session. This suggests that VPA had no effect on extinction learning. A previous human study (Kuriyama et al., 2011) also found that 400 mg of VPA had no effect on extinction learning, as quantified by SCL in a 3-day fear-learning paradigm. Furthermore, in that study, VPA had no effect on recall after extinction in a 3-day fear-learning paradigm, but it did show a reduced reinstatement of the conditioned association. Moreover, a study in mice showed that VPA enhanced memory consolidation for both the acquisition and the extinction of cued fear (Bredy and Barad, 2008). It is thus possible that the beneficial effects of VPA on subjective fear in the present study were related to an enhancement of the consolidation of extinction memories.

Prior studies have established VPA as a strong HDAC inhibitor (Bredy and Barad, 2008). However, it is important to note that our study did not specifically examine changes in histone acetylation. Consequently, we lack information as to whether VPA altered memory traces to transition them into a transcriptionally more permissive chromatin state, as has been previously shown in mice (Bredy and Barad, 2008; Gräff et al., 2014). If this occurred, it could have potentially served as a molecular mechanism behind the observed effects in the reduction of fear in the self-report questionnaires. Furthermore, VPA does not target class I HDACs with high affinity (Burns and Gräff, 2021) like HDAC2, which has been identified to mediate memory updating during extinction training in mice (Gräff et al., 2014). Thus, exploring whether alterations in histone acetylation and/or HDAC2 activity or whether more specific HDAC2-targeting HDACis mediate fear reduction following intervention and treatment could provide valuable insights for more refined studies in the future.

Limitations: Despite the large number of secondary outcome measures and the need for corrections for multiple comparisons, our study was based on a relatively small clinical sample size and was only sufficiently powered for our primary outcome. Furthermore, we did not have VPA and placebo groups without retrieval prior to exposure therapy as we had originally planned because it surpassed our resources due to slow recruitment. We therefore do not know if the same results would have been achieved without prior retrieval, based on the enhancing effect of VPA on the consolidation of extinction memories. Furthermore, one might argue that even with the inclusion of a control condition without retrieval, we would not be able to infer that our results are due to targeting the reconsolidation window, as the mere presence of the retrieval cue may have been sufficient to pharmacologically augment the extinction process. We are thus not able to verify all the criteria that have been suggested to be necessary for inferring reconsolidation was involved in the favorable effects of our intervention (for an overview, see Schroyens et al., 2023). Nevertheless, a study by Shiban et al. (2015) used a similar translation of the retrieval-extinction paradigm into a clinical application without any additional pharmacological interventions and included such a control group, and it found no improvement from retrieval before extinction-based therapy compared to standard extinction-based therapy without prior retrieval.

To conclude, our results indicate that a single administration of VPA before the retrieval of fear memories prior to exposure therapy led to a significant reduction in subjective fear levels 3 months later in comparison to the effects observed with no active substance and fear-memory retrieval prior to exposure therapy. This finding implies that the addition of VPA may hold promise as an additional therapeutic approach for enhancing treatment outcomes in individuals with spider phobia. However, the observed reduction in subjective fear did not translate into significant differences in objective behavioral or psychophysiological measures, indicating a need for a more comprehensive understanding of the complex interplay between subjective and objective outcomes in the treatment of phobias.

Future research should investigate the optimal dosage and frequency of VPA administration, the potential for variations in histone acetylation as a mediating factor, and the effects without retrieval. These considerations may contribute to a more comprehensive understanding of the mechanisms and therapeutic potential of VPA and HDAC inhibition in the treatment of specific phobias and potentially of other disorders treated with exposure therapy, which could ultimately improve treatment efficacy and patient outcomes.

## Contributors

DB, JG, and DQ designed the trial. DB, NS, DQ, and AZ drafted the paper. DB and NS supervised the data collection. JH, GI, MS, AZ, and DB collected the data. FD and CH contributed medical expertise. FD, CH, JH, UL, MS, and AZ commented on the design of the trial. NS and DB analyzed the data. All the authors commented on the paper.

## Data Availability

All data produced in the present study are available upon reasonable request to the authors.

## Acknowledgments

We thank Caroline Nicola Beer, Sandra Jenne, Sonu Sabnis for the data collection and for preprocessing the psychophysiological data, Michael Liedlgruber for assisting with the analysis of the psychophysiological data, Frodo Muijzer for technical advice on psychophysiological recordings, and Mathias Müller for programming the VR paradigm.

## References

1. Alberini, C.M., 2011. The Role of Reconsolidation and the Dynamic Process of Long-Term Memory Formation and Storage. Front. Behav. Neurosci. 5. 10.3389/fnbeh.2011.00012

2. American Psychiatric Association, 2013. Diagnostic and Statistical Manual of Mental Disorders, Fifth Edition. ed. American Psychiatric Association. 10.1176/appi.books.9780890425596

3. Arntz, A., Lavy, E., Van Den Berg, G., Van Rijsoort, S., 1993. Negative beliefs of spider phobics: A psychometric evaluation of the spider phobia beliefs questionnaire. Advances in Behaviour Research and Therapy 15, 257–277. 10.1016/0146-6402(93)90012-Q

4. Astill Wright, L., Sijbrandij, M., Sinnerton, R., Lewis, C., Roberts, N.P., Bisson, J.I., 2019. Pharmacological prevention and early treatment of post-traumatic stress disorder and acute stress disorder: a systematic review and meta-analysis. Transl Psychiatry 9, 334. 10.1038/s41398-019-0673-5

5. Auber, A., Tedesco, V., Jones, C.E., Monfils, M.-H., Chiamulera, C., 2013. Post-retrieval extinction as reconsolidation interference: methodological issues or boundary conditions? Psychopharmacology 226, 631–647. 10.1007/s00213-013-3004-1

6. Bentz, D., Michael, T., De Quervain, D.J.-F., Wilhelm, F.H., 2010. Enhancing exposure therapy for anxiety disorders with glucocorticoids: From basic mechanisms of emotional learning to clinical applications. Journal of Anxiety Disorders 24, 223–230. 10.1016/j.janxdis.2009.10.011

7. Bentz, D., Schiller, D., 2015. Threat processing: models and mechanisms. WIRES Cognitive Science 6, 427–439. 10.1002/wcs.1353

8. Bentz, D., Wang, N., Ibach, M.K., Schicktanz, N.S., Zimmer, A., Papassotiropoulos, A., de Quervain, D.J.F., 2021. Effectiveness of a stand-alone, smartphone-based virtual reality exposure app to reduce fear of heights in real-life: a randomized trial. npj Digit. Med. 4, 1–9. 10.1038/s41746-021-00387-7

9. Björkstrand, J., Agren, T., Åhs, F., Frick, A., Larsson, E.-M., Hjorth, O., Furmark, T., Fredrikson, M., 2017. Think twice, it’s all right: Long lasting effects of disrupted reconsolidation on brain and behavior in human long-term fear. Behavioural Brain Research 324, 125–129. 10.1016/j.bbr.2017.02.016

10. Bouton, M.E., 2004. Context and Behavioral Processes in Extinction: Table 1. Learn. Mem.11, 485–494. 10.1101/lm.78804

11. Bouton, M.E., 1993. Context, time, and memory retrieval in the interference paradigms of Pavlovian learning. Psychological Bulletin 114, 80–99. 10.1037/0033-2909.114.1.80

12. Bouton, M.E., Bolles, R.C., 1979. Role of conditioned contextual stimuli in reinstatement of extinguished fear. Journal of Experimental Psychology: Animal Behavior Processes 5, 368–378. 10.1037/0097-7403.5.4.368

13. Bredy, T.W., Barad, M., 2008. The histone deacetylase inhibitor valproic acid enhances acquisition, extinction, and reconsolidation of conditioned fear. Learn. Mem. 15, 39–45. 10.1101/lm.801108

14. Burns, A.M., Gräff, J., 2021. Cognitive epigenetic priming: leveraging histone acetylation for memory amelioration. Current Opinion in Neurobiology 67, 75–84. 10.1016/j.conb.2020.08.011

15. Chambless, D.L., Ollendick, T.H., 2001. Empirically Supported Psychological Interventions: Controversies and Evidence. Annu. Rev. Psychol. 52, 685–716. 10.1146/annurev.psych.52.1.685

16. Craske, M.G., Mystkowski, J.L., 2006. Exposure Therapy and Extinction: Clinical Studies., in: Craske, M.G., Hermans, D., Vansteenwegen, D. (Eds.), Fear and Learning: From Basic Processes to Clinical Implications. American Psychological Association, Washington, pp. 217–233. 10.1037/11474-011

17. De Quervain, D., Schwabe, L., Roozendaal, B., 2017. Stress, glucocorticoids and memory: implications for treating fear-related disorders. Nat Rev Neurosci 18, 7–19. 10.1038/nrn.2016.155

18. De Quervain, D.J.-F., Bentz, D., Michael, T., Bolt, O.C., Wiederhold, B.K., Margraf, J., Wilhelm, F.H., 2011. Glucocorticoids enhance extinction-based psychotherapy. Proc. Natl. Acad. Sci. U.S.A. 108, 6621–6625. 10.1073/pnas.1018214108

19. Elsey, J.W.B., Filmer, A.I., Galvin, H.R., Kurath, J.D., Vossoughi, L., Thomander, L.S., Zavodnik, M., Kindt, M., 2020. Reconsolidation-based treatment for fear of public speaking: a systematic pilot study using propranolol. Transl Psychiatry 10, 179. 10.1038/s41398-020-0857-z

20. Gräff, J., Joseph, N.F., Horn, M.E., Samiei, A., Meng, J., Seo, J., Rei, D., Bero, A.W., Phan, T.X., Wagner, F., Holson, E., Xu, J., Sun, J., Neve, R.L., Mach, R.H., Haggarty, S.J., Tsai, L.-H., 2014. Epigenetic Priming of Memory Updating during Reconsolidation to Attenuate Remote Fear Memories. Cell 156, 261–276. 10.1016/j.cell.2013.12.020

21. Hamlett, G.E., Foa, E.B., Brown, L.A., 2023. Exposure Therapy and Its Mechanisms, in: Milad, M.R., Norrholm, S.D. (Eds.), Fear Extinction, Current Topics in Behavioral Neurosciences. Springer International Publishing, Cham, pp. 273–288. 10.1007/7854_2023_428

22. Kindt, M., Elsey, J.W.B., 2023. A paradigm shift in the treatment of emotional memory disorders: Lessons from basic science. Brain Research Bulletin 192, 168–174. 10.1016/j.brainresbull.2022.11.019

23. Kindt, M., Soeter, M., Vervliet, B., 2009. Beyond extinction: erasing human fear responses and preventing the return of fear. Nat Neurosci 12, 256–258. 10.1038/nn.2271

24. Kuriyama, K., Honma, M., Soshi, T., Fujii, T., Kim, Y., 2011. Effect of d-cycloserine and valproic acid on the extinction of reinstated fear-conditioned responses and habituation of fear conditioning in healthy humans: a randomized controlled trial. Psychopharmacology 218, 589–597. 10.1007/s00213-011-2353-x

25. Lass-Hennemann, J., Michael, T., 2014. Endogenous cortisol levels influence exposure therapy in spider phobia. Behaviour Research and Therapy 60, 39–45. 10.1016/j.brat.2014.06.009

26. LeDoux, J.E., Pine, D.S., 2016. Using Neuroscience to Help Understand Fear and Anxiety: A Two-System Framework. AJP 173, 1083–1093. 10.1176/appi.ajp.2016.16030353

27. Lüdecke, D., Ben-Shachar, M., Patil, I., Waggoner, P., Makowski, D., 2021. performance: An R Package for Assessment, Comparison and Testing of Statistical Models. JOSS 6, 3139. 10.21105/joss.03139

28. Maples-Keller, J.L., Price, M., Jovanovic, T., Norrholm, S.D., Odenat, L., Post, L., Zwiebach, L., Breazeale, K., Gross, R., Kim, S.-J., Rothbaum, B.O., 2017. Targeting memory reconsolidation to prevent the return of fear in patients with fear of flying: M APLES -K ELLER ET AL . Depress Anxiety 34, 610–620. 10.1002/da.22626

29. Michael, T., Zetsche, U., Margraf, J., 2007. Epidemiology of anxiety disorders. Psychiatry 6, 136–142. 10.1016/j.mppsy.2007.01.007

30. Milad, M.R., Quirk, G.J., 2012. Fear Extinction as a Model for Translational Neuroscience: Ten Years of Progress. Annu. Rev. Psychol. 63, 129–151. 10.1146/annurev.psych.121208.131631

31. Monfils, M.-H., Cowansage, K.K., Klann, E., LeDoux, J.E., 2009. Extinction-Reconsolidation Boundaries: Key to Persistent Attenuation of Fear Memories. Science 324, 951–955. 10.1126/science.1167975

32. Nader, K., Schafe, G.E., Le Doux, J.E., 2000. Fear memories require protein synthesis in the amygdala for reconsolidation after retrieval. Nature 406, 722–726. 10.1038/35021052

33. Przybyslawski, J., Sara, S.J., 1997. Reconsolidation of memory after its reactivation. Behavioural Brain Research 84, 241–246. 10.1016/S0166-4328(96)00153-2

34. R Core Team, 2021. R: A language and environment for statistical ## computing. R Foundation for Statistical Computing, Vienna, Austria. ## URL https://www.R-project.org/.

35. Rescorla, R.A., 2004. Spontaneous Recovery. Learn. Mem. 11, 501–509. 10.1101/lm.77504

36. Rescorla, R.A., Heth, C.D., 1975. Reinstatement of fear to an extinguished conditioned stimulus. J Exp Psychol Anim Behav Process 1, 88–96.

37. Schiller, D., Monfils, M.-H., Raio, C.M., Johnson, D.C., LeDoux, J.E., Phelps, E.A., 2010. Preventing the return of fear in humans using reconsolidation update mechanisms. Nature 463, 49–53. 10.1038/nature08637

38. Schroyens, N., Beckers, T., Luyten, L., 2023. Appraising reconsolidation theory and its empirical validation. Psychon Bull Rev 30, 450–463. 10.3758/s13423-022-02173-2

39. Schwabe, L., Nader, K., Pruessner, J.C., 2014. Reconsolidation of Human Memory: Brain Mechanisms and Clinical Relevance. Biological Psychiatry 76, 274–280. 10.1016/j.biopsych.2014.03.008

40. Shiban, Y., Brütting, J., Pauli, P., Mühlberger, A., 2015. Fear reactivation prior to exposure therapy: Does it facilitate the effects of VR exposure in a randomized clinical sample? Journal of Behavior Therapy and Experimental Psychiatry 46, 133–140. 10.1016/j.jbtep.2014.09.009

41. Soeter, M., Kindt, M., 2015. An Abrupt Transformation of Phobic Behavior After a Post-Retrieval Amnesic Agent. Biological Psychiatry 78, 880–886. 10.1016/j.biopsych.2015.04.006

42. Spielberger, C.D., Gorsuch, R. L., Lushene, R., Vagg, P. R., Jacobs, G. A., 1970. Manual for the State-Trait Anxiety Inventory. Consulting Psychologists Press, Palo Alto, CA.

43. Steenen, S.A., Van Wijk, A.J., Van Der Heijden, G.J., Van Westrhenen, R., De Lange, J., De Jongh, A., 2016. Propranolol for the treatment of anxiety disorders: Systematic review and meta-analysis. J Psychopharmacol 30, 128–139. 10.1177/0269881115612236

44. Szymanski, J., O’Donohue, W., 1995. Fear of Spiders Questionnaire. Journal of Behavior Therapy and Experimental Psychiatry 26, 31–34. 10.1016/0005-7916(94)00072-T

45. Wolpe, J., 2018. Subjective Units of Distress Scale. 10.1037/t05183-000 Zimmer, A., Wang, N., Ibach, M.K., Fehlmann, B., Schicktanz, N.S., Bentz, D., Michael, T., Papassotiropoulos, A., de Quervain, D.J.F., 2021. Effectiveness of a smartphone-based, augmented reality exposure app to reduce fear of spiders in real-life: A randomized controlled trial. Journal of Anxiety Disorders 82, 102442. 10.1016/j.janxdis.2021.102442

