## Supplementary information for "Effects of the Histone Deacetylase Inhibitor Valproic Acid in Combination with Fear Memory Retrieval before Exposure Therapy for Spider Phobia: A Randomized Controlled Trial"

#### Amendments to the study protocol

The study protocol (version 1, 11-October-2015) was approved with requirements on December 16, 2015, by the Ethics Committee of Northwest and Central Switzerland (EKNZ), which resulted in study protocol version 2, 21-January-2016, and version 3, 10-March-2016. Swissmedic approved the study protocol (version 1, 11-October-2015) on January 11, 2016.

The study protocol foresaw the recruitment of 100 participants with a specific phobia (type: fear of spiders), 25 participants for each of the four planned groups: 1) intervention: valproic acid (VPA) + retrieval, 2) control I: placebo + retrieval, 3) control II: VPA + no retrieval, and 4) control group III: placebo + no retrieval. To ensure that the main research question of the study could still be addressed in the event of recruitment problems, the study protocol called for participants to be first recruited for the intervention group and control group I.

Because we were only able to recruit 46 participants within the first year of recruitment, we decided to conduct an interim analysis with the participants of the intervention group and control group I once 25 participants in each group had completed visit 3. The conduction of the interim analysis led to protocol version 4, 19-July-2017. In the end, 48 participants were included in the interim analysis (one participant was not able to attend visit 3 in the time frame set out by the study protocol, and another participant had to be excluded because of technical problems at visit 2). Importantly, no other changes to the statistical plan were made, especially not to the outcomes of the study. All changes to the protocol were approved by the EKNZ before the participants' group assignments were unblinded. The study was terminated after the interim analysis on May 24, 2018, as it did not support our primary hypothesis that VPA + retrieval would lead to reduced behavioral avoidance measured in the behavioral approach test (BAT) compared to placebo + retrieval. The participants that were already tested for control group II ($n$ = 7) and control group III ($n$ = 5) at that point of time were not analyzed.

#### Further information on methods and materials

#### Participants

Participants were recruited from the German-speaking general population of Switzerland using print, radio, and online advertisements. Participants were recruited for the study between August 15, 2016, and May 24, 2018. We included physically healthy individuals with DSM-IV specific phobia (animal type: spiders). We excluded individuals who were not fluent in German, had another DSM-IV axis-I disorder except further comorbid phobic disorders, were receiving concurrent psycho- or pharmacotherapy, had previously undertaken a therapy for their specific phobia with an exposure-based approach or were participating at the same time in another study, had a chronic medication intake, had a body weight less than 50 kg, had a history of coagulation or gastrointestinal disease or kinetosis, had abnormalities on physical examination by a medical doctor, showed clinically relevant deviations in laboratory values—blood count (including erythrocytes, leukocytes, thrombocytes, hemoglobin, and hematocrit), blood chemistry (including liver and pancreatic enzymes, kidney values, electrolytes, kidney values with creatinine and urea, albumin levels, vitamin B12, and TSH as thyroid function parameters)—from the normal range, were not normotensive (90/60–140/90 mmHg), smoked more than five cigarettes per day, or were not able to be abstinent from nicotine for at least 5 hours. We also excluded females who were pregnant or breastfeeding or who were not taking a birth-control pill or were not willing to implement a double-barrier birth-control method (because of the teratogenic effect of valproic acid). Body temperature was measured at the beginning of each study day to exclude acute infective diseases, and participants were rescheduled if their body temperature was >38°C. On the day of visit 2, exclusion criteria were examined again, the body temperature and vital signs of the participants were taken, and all the participants had to provide a urine sample for a drug screening for the following substances: amphetamines, benzodiazepine, cocaine, methamphetamine, morphine/opium, and THC (Stephany

Diagnostika). This test had to be negative for study continuation. For females an additional pregnancy urine test was conducted.

**Study setting**

The study was conducted in two laboratory rooms at the University Psychiatric Clinics Basel. One room was used for clinical assessments, filling in questionnaires, medication intake, waiting after medication intake, and for viewing a film after the retrieval procedure at visit 2. The second room was dedicated to exposure in virtual reality (VR), BATs, and psychophysiological measurements. Exposure and BAT in VR took place while the experimental room was darkened with opaque blinds. Temperature in the experimental room was monitored with a LogTag Temperature Data Logger (TRID30-7) to ensure consistent psychophysiological measurements (target range: 15–25°C).

**Procedure**

Potential participants received detailed information via email, and additional information, including inclusion and exclusion criteria, by phone. Subsequently, eligible individuals were invited for an in-person screening at the University Psychiatric Clinics Basel, Switzerland, where all three study visits took place.

At the beginning of visit 1, written informed consent was obtained. Information about the socioeconomic status of the potential participants was queried. An examination of medical status, medication history, a blood sample, and a physical examination were undertaken by a medical doctor. Then, a structured clinical interview (DIPS DSM-IV-TR) (Schneider and Margraf, 2011) was conducted by a trained clinical rater to validate the diagnosis of a specific phobia (animal type: spider) and to exclude patients with comorbid mental disorders other than a further specific phobia. Next, visual analogue scales (VAS) were used to assess the participants' motivation and possible adverse events (AEs), and vital signs (heart rate; HR, blood pressure; BP) were taken. Furthermore, we assessed the potential participants' depressive symptoms with the Beck's Depression Inventory (BDI-II) (Beck et al., 2011), their mental state with a multidimensional questionnaire (MDBF-A/B) (Steyer et al., 2017), and their state and trait anxiety with the State-Trait Anxiety Inventory (STAI-S/T) (Laux, L et al., 1981; Spielberger, Charles D. et al., 1970). Self-reported fear of spiders was measured using the German version of the Fear of Spiders Questionnaire (FSQ) (Rinck et al., 2002; Szymanski and O'Donohue, 1995) and the Spider Phobia Beliefs Questionnaire (SBQ) (Arntz et al., 1993; Pössel and Hautzinger, 2003), and symptoms of simulation sickness using the Simulation Sickness Questionnaire (SSQ) to obtain a baseline measure before VR immersion for the queried symptoms (Kennedy et al., 1993). Afterward during the phobic-picture-stimuli task (PPST), psychophysiological (electrodermal activity; EDA, electrocardiography; ECG, electromyography; EMG) and subjective reactions (valence, arousal, anxiety) to neutral, negative, and fear-related stimuli were measured during the PPST. A BAT was conducted in VR, while EDA and ECG were continuously measured, and a Subjective Units of Distress Scale (SUDS) (Wolpe, 2018) was queried. This was followed by a second SSQ and the Igroup Presence Questionnaire (IPQ) (Schubert, 2003) was used to assess presence in VR. The BAT in vivo was conducted and SUDS measurements were taken. At the end of visit 1, vital signs were taken, and the participants completed a VAS and received psychoeducational material to understand the rationale of exposure therapy. Therapy expectations were collected via credibility or expectancy-for-improvement scales by Borkovec and Nau (1972), which were adapted for the purposes of the study.

Visit 2 took place between 7 and 21 days after visit 1. At the beginning of visit 2, the major inclusion and exclusion criteria were rechecked, and each participant's health status was assessed, including a pregnancy test for females and drug screening for all the participants. Afterward, the participants completed a VAS to assess possible AEs and motivation, and vital signs were taken. Participants filled out the MDBF and STAI-S. A blood sample was taken to assess VPA blood levels at baseline. Participants were then given a single oral dose of 8.3 ml VPA syrup (Orfiril ®, active pharmaceutical ingredient: 500 mg VPA) or 8.3 ml placebo syrup. Participants had to wait for 3 hours and 10 minutes after drug intake before starting exposure therapy in VR (resp. 3 hours before retrieval). During the waiting period, participants completed a VAS to assess both possible AEs and motivation, and vital signs were taken every 30 minutes. At the end of the waiting period, participants filled out an SSQ and indicated their perception about whether they received VPA or a placebo on an additional VAS (VAS VPA or placebo intake). Then, the retrieval procedure was performed followed by an SUDS measurement. Afterward, they had to see a 10-minute-long documentary of neutral content before the begin of exposure therapy in VR. Afterward, participants filled out another SSQ and an IPQ. VAS was used to assess possible AEs, and vital signs were taken. Participants filled out a second MDBF, STAI-S, as well as the FSQ, and SBQ. Additionally, they were asked a second time about their perception about whether they received VPA or a placebo on a VAS. A second blood sample was taken to assess VPA blood levels. The VAS was used to assess possible AEs, and vital signs were taken. Forty-eight hours after visit 2, participants received a link via email to indicate their intake of medication, drugs, and alcohol since visit 2 to control for negative effects on memory formation.

Visit 3 was scheduled 90±10 days after the exposure therapy in VR at visit 2, within the time frame November 28, 2016, to February 17, 2018. Participants' body temperature and vital signs were taken and they filled out a VAS to assess possible AEs and motivation. All participants underwent the same drug screening as at visit 2. Additionally, participants had to fill out the FSQ, SBQ, SSQ, and a questionnaire about whether self-exposure to spiders had occurred between visit 2 and 3. Then the existence of the DSM-IV diagnosis for a specific phobia (animal type: spiders) was reevaluated by a trained clinical rater using the DIPS (Schneider and Margraf, 2011). Just as at visit 1, the PPST, a BAT in VR and in vivo were conducted. Additionally, a second BAT in VR in an unfamiliar virtual room was conducted. Afterward, participants filled out a second SSQ and an IPQ and were asked a last time about their perception of whether they received VPA or a placebo. Furthermore, a pictorial n-back task (Loos et al., 2020) and a picture-recognition task were conducted to explore the potential effects of working memory load on immediate fear of spiders regardless of group assignment (pilot data for another study, data is not included in the report). Prior to the participants' dismissal, inquiries regarding potential AEs were conducted, and their vital signs were assessed.

### Study tasks

#### BAT in vivo

The BAT in vivo procedure was similar to the one used by Lass-Hennemann and Michael (2014) (2014). Participants were guided to the closed door of the experimental room and were instructed to approach a living house spider (Tegenaria atrica) measuring about 5 cm placed in a sealed transparent plastic container (size: 30 × 20 × 15 cm) at the far end of the room. Participants were instructed to approach the spider in the following manner: to open the door, approach the plastic container, touch the container, remove the lid of the container, put a hand in the container, touch the spider with a forefinger, and hold the spider for at least 20 s. Participants were instructed that the BAT served as a measurement of their current approach behavior and that they should not force themselves to do something they would not do under normal circumstances. The BAT in vivo was terminated once a participant indicated not being able to proceed any further during the BAT, once the last step was accomplished, or after 3 min, the maximum time for accomplishing the BAT in vivo. Each step in the task completed by the participant translated into a score from 0 to 12, 0 = refused to enter the test room, 1 = stopped 5 m away from the container, 2 = stopped 4 m away from the container, 3 = stopped 3 m away from the container, 4 = stopped 2 m away from the container, 5 = stopped 1 m away from the container, 6 = stopped close to the windowsill with the container, 7 = touched the container, 8 = removed the lid, 9 = put a hand in the container, 10 = touched the spider with one forefinger, 11 = held the spider for less than 20 s, and 12 = held the spider for at least 20 s. The BAT in vivo was terminated once the participant indicated not being able to perform further steps, once the last step was accomplished, or after a maximum of 3 minutes. After the BAT was terminated, participants were asked with an SUDS about their fear (0 = no fear, 10 = maximum fear). Before starting the BAT in vivo, participants were asked how fearful they expected they would be during the BAT in vivo with an SUDS (0 = no fear, 10 = maximum fear imaginable).

**Retrieval procedure before exposure therapy**
Participants were shown a virtual spider for 5 s in a virtual room, which was modeled after the actual experimental room (see Figure 2) to initiate retrieval of fear memories. This was done 10 minutes prior to exposure therapy in VR to target the reconsolidation window (Schiller et al., 2010). Participants were told that a spider might be moving toward them to reinforce fear-memory reactivation, even though the spider did not move. Afterward participants were asked to indicate the fear they experienced during the presentation of the virtual spider via an SUDS. Electrodermal activity and ECG were measured during the 5 s of the presentation of the virtual spider.

**Exposure therapy**
Our VR exposure paradigm resembled the exposure protocol employed in the study by Shiban et al. (2015). After the instructions were given, exposure in VR started. The exposure session consisted of ten standardized VR exposure scenes with increasing difficulty (hierarchical exposure therapy approach), each scene lasting for 3 min (total duration of the exposure: 30 min). Participants were presented with a single spider in the first scene, and this built up to four different spiders at once, crawling around, during the last scenes. The virtual room where the spiders were presented was modeled after the actual experimental room. VR exposure took place while the participants were seated; the experimenter monitored the session on a PC screen. Instructions and fear assessments via an SUDS were provided through headphones and displayed in VR.
To assess within-session fear reduction, participants were exposed to the first scene with only one spider again at the end of the exposure session (see also SF2). Five seconds after the start of each scene and 15 s before the end, fear was measured with an SUDS (0 = no fear and 10 = maximum fear). Participants could exit the exposure in VR at any time, though they were discouraged from doing so by being told that this would not be therapeutically advised. Participants were instructed to focus on the spider, the triggered emotions, bodily experiences, and fear-related cognitions and to avoid cognitive avoidance strategies.

**BAT in VR**
BAT in VR took place with participants seated; the experimenter monitored the session on a PC screen. Instructions and fear assessments using the SUDS were provided through headphones and displayed in VR. At the beginning of the BAT in VR, participants were situated in a virtual corridor that was modeled after the corridor leading to the actual experimental room. In the virtual corridor, participants first practiced how to navigate in the VR scenario with the provided gamepad. Then they received the instruction over headphones and displayed in VR that they would soon be standing in a laboratory room with a spider on the opposite wall and that their task was to approach the spider. They were instructed that they would have a maximum time of 3 min to approach the spider. Participants were told that if they were not willing to approach closer to the spider, they should remain as close as possible to it for the remaining time. After the instructions, participants were asked to indicate via an SUDS how fearful they thought they would become while executing the BAT in VR. Once participants

indicated that they were ready to start the BAT in VR, they entered the virtual experimental room with the virtual spider. For the first 15 s, participants were not able to move and were asked after this amount of time passed about their fear via an SUDS. After that, they could move freely in any direction in the virtual room using the gamepad and approach the spider. As the virtual room was modeled after the actual experimental room, the distance from the door to the virtual spider at the end of the room could be translated to 5 m in real life. At the end of the 3 min, participants were asked to indicate their fear another time via an SUDS. The second BAT in VR (BAT in VR new context) was designed to ease the possible return of fear at visit 3. The sequence resembled the BAT described above; the only difference was that the BAT took place in an unfamiliar virtual room.

**Phobic-picture-stimuli task (PPST)**

The phobic-picture-stimuli task (PPST) consists of fear-related (spider), negative (snake), and neutral pictures. Pictures of all three categories were taken from IAPS or GAPED (Bradley, M. M., & Lang, P. J, 2007; Dan-Glauser and Scherer, 2011). Psychophysiological measurements (electrodermal activity/EDA, electrocardiography/ECG, and electromyography/EMG) were taken continuously during the PPST. During the PPST, participants were comfortably seated on a chair at a distance of 50 cm to the screen on which the PPST was presented. Participants were verbally instructed to sit still and relax and look at the computer screen at all times during the PPST, even when no pictures and only a white cross were shown. Participants were instructed to keep their hand on which the electrodes for EDA measurements were attached still but not to stiffen it. All further instructions for the PPST were given in writing on the computer screen. The PPST started by measuring participants' physiological baseline (duration: 3 min), during which participants were instructed to sit still but to be relaxed. After the physiological baseline, the training phase for the main phase of the PPST began. The training phase was followed by a startle-habituation phase. After the startle-habituation phase, the main phase of the PPST started. The duration of the main phase of the PPST was approximately 20 min.

The phases of the PPST were composed of the following. During the training phase, participants became accustomed to the sequence of the PPST and the subjective rating format used during the main phase of the PPST. During the training phase, three neutral pictures were shown (duration: 6 s each) that participants had to rate for valence (left anchor: negative; right anchor: positive), arousal (left anchor: agitating/unsettling; right anchor: calm), and anxiety (left anchor: very big; right anchor: not at all) on a visual analogue scale (VAS). Participants had to click with the mouse on the horizontal VAS to rate the elicited valence, arousal, or anxiety after looking at the picture. There was no time limit for giving the response. The rating positions on the VAS were transformed to a scale from 0 (left anchor) to 100 (right anchor). To facilitate the interpretability of the polarity of the valence, the ratings were reversed. Scores >50 refer to negative valence, high arousal, and high anxiety; 50 refers to neutral valence, arousal, and anxiety; and scores <50 indicate positive valence, low arousal, and low anxiety. During the presentation of the first picture, a startle tone was played as well during the intertrial interval (ITI) of the third picture. Participants were told that tones would be played over the headphones and that they should not be bothered by that. During the startle-habituation phase participants were instructed to sit still and fixate on a cross on the screen for 2 min while five startle probes (white noise, 100 dB) were presented. Participants were told that a tone would be played over the headphones. During the main phase, 24 pictures (eight pictures per category: spider, snake, neutral) were presented for a duration of 6 s each, followed by an ITI of 15,000–20,000 ms before the VASs for valence, arousal, and anxiety described above were presented. During the main phase, 24 startle probes were presented: 12 startle probes were presented 3000–4500 ms after the start of each picture presentation (four on each picture category), and 12 during the ITIs at random 3000–4500 ms after the start of the ITI. Twelve pictures (four from each category) were presented without a startle probe on the picture or the following ITI. Participants were instructed that startle probes might be presented that the participants could ignore during the main phase of the PPST. The sequence of pictures of the main phase was quasi-randomized, with no more than three pictures from the same category in a row and no more than three startle conditions in a row.

### Questionnaires

**Spider phobia:** Fear of spiders was assessed using the German version of the Fear of Spiders Questionnaire (FSQ, German version: FAS) (Rinck et al., 2002; Szymanski and O'Donohue, 1995) and the Spider Phobia Beliefs Questionnaire (SBQ) (Arntz et al., 1993; Pössel and Hautzinger, 2003). The FSQ is a self-report questionnaire and measures avoidance behavior as well as the fear of harm. It consists of 18 items rated on a 7-point scale (0 = not at all true to 6 = very true, $\alpha = 0.96$, range 0–108), higher scores indicating greater severity. The SBQ is a self-report questionnaire and specifically assesses spider-related catastrophic cognitions and emotions. It consists of 48 items (0% = I do not believe that at all to 100% = I am absolutely convinced, $\alpha = 0.98$) separated into spider-related and self-related beliefs. A mean score can be calculated over all items of both subscales (range 0–100). Higher scores indicate greater severity. Self-reported change in participants' fear of spiders was queries with a VAS (10 cm long, left anchor: not at all (0), right anchor: very strong (100)).

**Mood, anxiety:** The BDI-II (Beck et al., 2011) is a self-report questionnaire assessing depressive symptoms. It consists of 21 items, each of which is scored on a scale from 0 to 3 (range 0–63). Higher values indicate more severe symptoms (0–8: no depression, 9–13: minimal depression, 14–19: mild depression, 20–28 moderate depression, 29–63: severe depression). Participants who scored 13 or higher on BDI-II or answered the question about suicidal tendencies with a score of 1 or higher were referred to a member of the study team with clinical training, and excluded from further participation in the study. The multidimensional mood questionnaire (German version: Mehrdimensionale Befindlichkeitsfragebogen, MDBF) consists of a 24-item-long version; items are rated on a 5-point scale to measure three bipolar dimensions of the current psychological situation: good–bad mood, wakefulness–tiredness, and calmness–restlessness. The MDBF long version can be separated in two parallel short versions (MDBF-A and B) with 12 items each (3 items for each dimension; range 8–40) (Steyer et al., 2017). The State-Trait Anxiety Inventory (STAI) is a self-report questionnaire and measures trait (STAI-T) and state (STAI-S) anxiety (Laux, L et al., 1981; Spielberger, Charles D. et al., 1970). It consists of 20 items assessing trait and state anxiety on a 4-point scale. The trait and state sum score ranges between 20 and 80. Higher scores indicate greater anxiety.

**VR:** Presence in VR was measured with the German version of the Igroup Presence Questionnaire (IPQ) (Schubert, 2003). The IPQ consists of 14 items (range −42–42, with higher scores corresponding to a stronger feeling of presence) parted into three subscales for spatial presence (five items), involvement (four items), and experienced realism (four items) as well as one item for general presence. The Simulator Sickness Questionnaire (SSQ) was implemented as a safety measure to assess side effects of VR immersion during the BAT in VR at visit 1 and 3 as well as during the exposure in VR at visit 2. The SSQ consists of 16 items indexing common side effects of VR immersion, e.g., headache, nausea, or sweating. The items are rated on a 4-point scale (from 0 = none to 3 = very strong), resulting in a total score with a range of 0–48, higher scores indicating greater severity. The SSQ was given two times during visits 1 and 3: once at baseline before any outcomes were assessed and once directly after VR immersion during the BAT in VR; during visit 2, it was given once directly before the retrieval procedure and a second time directly after the VR exposure.

**Treatment:** Treatment expectations were measured with a version of the credibility/expectancy-for-improvement scales by Borkovec and Nau (1972) that had been adapted for our study's purposes (treating a fear of spiders): the scale consists of five items on expectations of improvement from treatment on a 10-point scale (0 = not at all to 10 = very much, α = 0.81, range 0–50). Participants were asked using a VAS (10 cm long, left side: placebo, right side: medication) whether they believed they had received a placebo or VPA directly before and directly after exposure therapy at visit 2 and at the end of visit 3.

**Safety measures:** To assess adverse events (AEs) of the study medication and the experimental procedure in general, we regularly assessed the symptoms of headache, stomach pain, nausea, fatigue, dizziness, drowsiness, muscle fatigue, and motivation using horizontal VASs and measured participants' vital signs (blood pressure/BP and heart rate/HR). The anchors of the 10 cm long VAS were on the left side "not at all" (0) and the right side "very strong" (100). VAS values >45 (values on the VAS motivation were not considered) and BP measurements outside the range 90/60 and 140/90 mmHg were considered critical and were followed up by a medical doctor from the team. The VAS and vital signs were taken at baseline and at the end of visits 1 and 3 and at visit 2 at baseline, drug intake +30, +60, +90, +120, +150, after VR exposure, and at the end of visit 2 (for more information, see SF3 and ST4).

In each case, the German version of the questionnaire was used. All questionnaire data was collected via SoSci Survey (Version 2.600-i) (Leiner, 2019).

### Technical information on the material for the experimental tasks

#### VR setup

The VR paradigms for the exposure therapy and BAT in VR were developed by the company VT+ GmbH (Würzburg, Germany). VT+ GmbH also provided the setup for VR delivery, which consisted of a Windows PC with a Windows 7 Professional operating system to render and control the VR paradigm, a built-in iPad with iOS 10.2 (14C92) interface to control the VR paradigm, a 24-inch screen (1920 × 1080) for the experimenter to follow the VR session, and a second Windows PC (Intel i7-4702HQ 2.2GHz) with a 24-inch screen (1920 × 1080) to record and control the psychophysiological measurements. The integration of the devices for the psychophysiological measurements is described in detail below. An Oculus Rift DevKit-2 (Oculus VR Inc., Menlo Park, California, USA) with a resolution of 960 × 1080 pixels per eye and 100° FOV-D was used as Head Mounted Display (HMD). The Logitech Gamepad F310 (Apples, Switzerland) was used for movement in VR. For the delivery of audio signals, headphones from Beyerdynamic (Modell DT 770 M, Heilbronn, Germany) were used. The rendering of the virtual scenes was performed with the Source engine (Valve Inc., Bellevue, Washington, USA). The software CyberSession (CS Expo; Version 5.6.98-DEBUG) was used to control the VR environment. The experimenter controlled the CyberSession software via the built-in iPad via an iOS app (CS-Remote).

#### Setup for psychophysiological measurements

Physiological measurements (EDA, ECG, EMG) during the exposure therapy and BAT in VR as well as the PPST were recorded using a Porti 32-channel amplifier and the software Polybench (TMSi, Twente Medical Systems International, EJ Oldenzaal, Netherlands). Event markers generated by the experiment control script and the VR simulation were transmitted from the image-generator parallel port via TMSi Porti Interface using the TMCSi DC-Box with a Fusbi fiber-optic connector that was integrated into the VT + VR exposure system.

### Information on physiological measurements

#### Measuring electrodermal activity

Participants were instructed to wash their hands with soap. Skin-conductance level (SCL) was measured via two nondisposable Ag/AgCl electrodes filled with isotonic electrode paste. They were placed on the middle phalanx of the index and middle fingers of the left hand and secured with a velcro strap attached to the electrodes. The constant voltage between the electrodes was 0.5 V. Data was stored on a personal computer with a sampling frequency of 512 Hz (Fowles et al., 1981). SCLs were recorded continuously in microsiemens (mS). Afterward, the

Autonomic Nervous System Laboratory version 2.6 (ANLSAB) was used to visually inspect and edit artifacts (Blechert et al., 2016).

**Measuring electrocardial activity**

Cardiovascular activity was measured by electrocardiography (ECG) using three disposable 55 mm diameter Ag/AgCl solid-gel electrodes attached to alcohol-cleansed skin sites. One electrode was attached on the breast bone, one on the lower ribcage, and one ground electrode on the shoulder. The signals were digitized with a 1024 Hz sampling rate. Afterward, R-waves were determined automatically using the software ANSLAB (Blechert et al., 2016), followed by a visual inspection and manual editing for occasional misdetections, artifacts, and ectopic beats. HR was quantified in beats per min (bpm).

**Measuring electromuscular activity**

The EMG electrodes were placed below the right eye on the skin surface overlying the orbicularis oculi muscle. Facial muscular activity was assessed using two nondisposable 2 mm inner-diameter Ag/AgCl electrodes attached to alcohol-cleaned skin below the right eye that had been just peeled with Nuprep paste. The inner electrode was placed directly under the right pupil (when the participant looked straight) and the outer electrode 1–2 cm lateral to the inner electrode with a small gap. The electrodes were filled with a highly conductive electrode gel and attached with double-sided adhesive collars. The raw EMG signal was recorded with a sampling rate of 1000 Hz. EMG amplitude (in µV) was assessed as an index of muscular activity after signal filtering and rectification using established criteria (Blumenthal et al., 2005). ANSLAB (Blechert et al., 2016) was used for raw data filtering and extraction of startle responses. The startle EMG was 50 Hz notch filtered, rectified and the magnitude of the startle response (amplitude) in a response window of 20 and 150 milliseconds (ms) following the startle noise was computed in µV. Amplitude refers to the difference between the peak of the startle response and the baseline immediately preceding the eliciting-stimulus onset.

**Data reduction and statistical analysis of psychophysiological measurements**

For the **BAT in VR**, we extracted the mean SCL and HR for the 15 s after the start of BAT in VR when the participants entered the room and saw the spider at the opposite virtual window for the first time and while the participants were not able to move freely in the VR. For SCL we used only the interval between +1 and +15 s after the start of BAT in VR, for HR we used the complete interval between 0 and +15. For baseline correction we subtracted the mean SCL for 1 s (interval between −1 and 0) respectively for HR 3 s (interval between −3 and 0) before the participant entered the virtual experimental room with the virtual spider.

For **retrieval**, we extracted the mean SCL for the interval between +1 and +5 s after the presentation of the retrieval cue (virtual spider) and the mean HR for the complete interval between 0 and +5 s after the presentation of the retrieval cue. For baseline correction we subtracted the mean SCL for one second interval between −1 and 0) respectively for HR 3 s (interval between −3 and 0) before the presentation of the retrieval cue.

During **exposure therapy in VR**, we extracted the mean SCL and HR for the first and last 15 s of exposure therapy of each of the 10 exposure scenes (in accordance with Shiban et al. (2015) (2015) that used the same exposure paradigm). For SCL we only used the interval between +1 and +15 s for the first 15 s of each exposure scene, for HR and the last 15 s of each exposure scene we used the mean of the complete interval. For baseline correction we subtracted the mean SCL for 1 s (interval between −1 and 0) respectively for HR 3 s (interval between −3 and 0) before each exposure scene.

During **PPST**, we extracted the mean SCL for the interval between +1 and +6 s and HR for the complete 6 s of each nonstartled picture as well as the startle reaction to the startled pictures. For baseline correction we subtracted the mean SCL for 1 s (interval between −1 and 0) respectively for HR 3 s (interval between − 3 and 0) before each picture presentation. Means for SCL, HR, and startle amplitude were calculated for each picture category (spider, snake, neutral). Participants that indicated an additional specific phobia (animal type: snake) during the DIPS at visit 1 were excluded from this analysis.

**Secondary outcomes**
1) SUDS after BAT in vivo. The SUDS score ranged from 0 = no fear to 10 = maximum fear. 2) Approach behavior BAT in VR quantified by the virtual distance to the spider at the end of BAT in VR translated to meters in real-life (ranging from 0 to 5). 3) SUDS BAT in VR taken 15 s after participants entered the virtual room and saw the virtual spider at the opposite window for the first time. The score ranged from 0 = no fear to 10 = maximum fear. 4) HR BAT in VR based on the mean baseline-corrected HR during the first 15 s of the BAT in VR quantified in bpm. 5) SCL BAT in VR based on the mean baseline-corrected SCL during the time from 1 s after the start BAT to 15 s after the start BAT quantified in mS. 6) Valence rating of spider pictures based on the mean valence rating of nonstartled spider pictures during the PPST. 7) Arousal rating of spider pictures based on mean arousal rating of nonstartled spider pictures during the PPST. 8) Anxiety rating of spider pictures based on mean anxiety rating of nonstartled spider pictures during the PPST. The score of valence, arousal, and anxiety ratings of nonstartled pictures during the PPST ranged from 0 to 100; 100 refers to negative valence, high arousal, and high anxiety. 9) HR spider pictures based on mean baseline-corrected HR for the complete 6 s of each nonstartled spider picture of the PPST in bpm. 10) SCL spider pictures based on mean baseline-corrected SCL for the 1–6 s of each nonstartled spider picture of the PPST quantified in mS. 11) Startle spider pictures based on mean startle amplitude during the response window of 20 and 150 ms following the startle on the spider picture during the PPST quantified in microvolts (mV). 12) FSQ (sum score ranges from 0–108). 13) SBQ (mean sum score 0–100). 14) STAI-S (sum score ranges from 20–80). 15) DSM-IV impairment score based on the DIPS for DSM-IV-TR. 16) DSM-IV distress score based on the DIPS for DSM-IV-TR. DSM-IV impairment and distress score range from 0 to 8.

**Additional analysis**
Model assumptions were evaluated using the Performance package in R (Lüdecke et al., 2021). In case where assumptions such as normality, homoscedasticity, or collinearity were violated or influential data points (Cook's Distance ≥ 0.5) were identified, the dependent variables were residualized for the covariates age and sex. Subsequently, nonparametric Mann–Whitney tests for independent samples were applied to test for group differences. In the context, we employed the effect size measures *V*. Interpretation of the *V* effect size is as follows: a *V* value of <0.3 is considered small, between 0.3 and 0.5 is regarded medium, and >0.5 is classified as large.

**Participants' perception of intake of VPA or placebo**
Participants' perception of intake of VPA or placebo was measured with a VAS (0 = placebo, 100 = medication, valproic acid). After medication intake directly before the exposure at visit 2, nine out of 22 participants in the VPA + retrieval group (compared to 12 out of 26 participants in the placebo + retrieval group) indicated higher values than 50 on the VAS (0 = placebo, 100 = medication, VPA + retrieval: mean = 35.6, *SD* = 25.7; placebo + retrieval: mean = 34.8, *SD* = 28.3). After exposure at visit 2, nine out of 22 participants in the VPA + retrieval group (compared to 12 out of 26 participants in the placebo + retrieval group) indicated higher values than 50 on the VAS (0 = placebo, 100 = medication, VPA + retrieval: mean = 46.0, *SD* = 28.6; placebo + retrieval: mean = 45.0, *SD* = 31.6). At the end of visit 3 after performing the study tasks (including BAT), 11 out of 22 participants (one missing answer) in the VPA + retrieval group (compared to 10 out of 26 participants in the placebo + retrieval group) indicated higher

values than 50 on the VAS (0 = placebo, 100 = medication, VPA + retrieval: mean = 48.9, *SD* = 33.8, mean placebo + retrieval: mean = 35.2, *SD* = 34.7).

### Fear reduction over time

To investigate fear reduction over time, that is, to compare phobic fear at baseline and at follow-up, we ran additional nonparametric two-way ANOVAs by means of ANOVA-type statistic (ATS) as provided in the R package nparLD (Noguchi et al., 2012), a strategy not predefined in the study protocol. We opted for nonparametric models instead of linear-mixed models because the assumptions of the lme models were violated. Our dependent variables included both primary and secondary outcomes (assessed at visits 1 and 3, without calculating difference scores). The independent variables were group allocation and visit. To enable the use of nparLD models, we first residualized the primary and secondary outcomes for age and sex. Subsequently, these residualized variables were employed as dependent variables in the nonparametric two-way ANOVAs, with visit (within-subject variable) and group allocation (between-subject variable) as the independent variables. For FSQ ($F$ (1) = 9.84, $p$ = 0.00171) and SBQ ($F$ (1) = 9.91, $p$ = 0.00164) there was—as according to the main analyses—a significant interaction between visit and group allocation. Post hoc tests for each group separately revealed a significant reduction of phobic fear after exposure in both VPA + retrieval and placebo + retrieval for FSQ (VPA + retrieval: $F$ (1) = 132.19, $p$ = 1.36E-30; placebo + retrieval: $F$(1) = 24.03, $p$ = 9.47E-07) and SBQ (VPA + retrieval: $F$ (1) = 59.42, $p$ = 1.27E-14; placebo + retrieval: $F$(1) = 23.17, $p$ = 1.49E-06), but it was more pronounced for VPA + retrieval. For all the other dependent variables, there were no significant interaction effects (all $p \geq 0.0668$), nor main effects of group allocation (all $p \geq 0.069$). A significant main effect of time was detected for the primary outcome approach behavior in BAT in vivo ($F$(1) = 40.73, $p$ = 0.00164), and for the secondary outcomes SUDS after BAT in vivo ($F$(1) = 33.46, $p$ = 7.27E-9), performance BAT in VR ($F$(1) = 94.88, $p$ = 2.02E-22), SUDS BAT in VR ($F$(1) = 141.97, $p$ = 9.89E-33), SCL BAT in VR ($F$(1) = 13.78, $p$ = 2.06E-4), arousal rating spider pictures ($F$(1) = 18.34, $p$ =1.85E-5) and anxiety rating spider pictures ($F$(1)=26.65, $p$ =2.44E-7) during the PPST, startle spider pictures ($F$(1) = 9.82, $p$ = 1.73E-3) and HR spider pictures ($F$(1) = 11.45, $p$ = 7.16E-4) during the PPST, DSM-IV impairment ($F$(1) = 21.39, $p$ = 3.74E-6), and DSM-IV distress ($F$(1) = 56.10, $p$ = 6.87E-14). No significant effect of time was found on valence rating spider pictures during the PPST ($F$(1) = 8.71, $p$ = 0.003167), HR BAT in VR ($F$ (1) = 4.34, $p$ = 0.04), and SCL spider pictures during the PPST ($F$(1) = 2.96, $p$ = 0.09), and the STAI-S ($F$(1) = 0.94, $p$ = 0.33) (for more information, see also ST1-3 and SF1).

### Fear reduction during exposure in VR

SF4 illustrates the SUDS ratings, HR, and SCL for both groups during exposure in VR at the beginning and end of all 10 scenes. To examine potential group differences in the SUDS ratings, HR, and SCL, dependent variables were residualized for age and sex, and then the nonparametric ANOVA by means of ANOVA-type statistic (ATS), as provided in the R package nparLD (Noguchi et al., 2012), was used by including group, scene, and time point as independent variables. The ATS rank-based method tests the hypothesis of equality of distributions rather than the equality of means (Shah and Madden, 2004).

**SUDS:** No significant three-way interaction between time point, scene, and group was found ($p$ = 0.79). However, there was a nominal significant two-way interaction between scene and group (*WTS* (3.9) = 2.78, $p$ = 0.026), as well as a significant two-way interaction between time point and scene (*WTS* (6.34) = 16.78, $p$ = 1.98E-20). Post hoc tests for each scene separately revealed nominal significantly higher SUDS ratings in the VPA + retrieval group compared to the placebo + retrieval group for scene 1 and 4 (scene 1: *WTS* (1) = 13.62, $p$ = 0.00022, scene 4: *WTS* (1) = 5.55, $p$ = 0.0184). Furthermore, in all the scenes, there was a significant main

effect of time point, indicating a decrease in SUDS ratings from the beginning to the end of each scene (all $WTS > 19.63$, all $p < 9.38E-06$).
Scene 1 and 10 were identical; consequently, we reran the analyses, this time including only scene 1 and 10, to determine if a significant decrease occurred. Comparing the SUDS ratings in scene 1 to those in scene 10, no statistically significant three-way interaction was observed among timepoint, scene, and group allocation ($p = 0.88$), nor were there significant two-way interactions between group and timepoint or between time point and scene (all $p > 0.70$). However, a significant two-way interaction between group and scene on the SUDS (*ATS* (1) = 13.46, $p = 2.44E-04$). Subsequent post-hoc tests, conducted separately for both groups, demonstrated a significant main effect of time point in both the placebo + retrieval (*ATS* (1) = 21.39, $p = 3.75E-06$) and the VPA + retrieval group (*ATS* (1) = 29.35, $p = 6.05E-08$), as well as a significant main effect of scene in the placebo + retrieval group (*ATS* (1) =84.85, $p = 3.22E-20$) and the VPA + retrieval group (*ATS* (1) = 110.81, $p = 6.51E-26$). These findings suggest a more pronounced reduction in SUDS ratings from level 1 to level 10 in the VPA group. Comparing SUDS ratings in scene 1 timepoint beginning and scene 10 timepoint end of the scene, no significant two-way interaction was observed between group and timepoint (*ATS* (1) = 2.52, $p = 0.11$), and no significant main effect of group (*ATS* (1) = 0.57, $p = 0.45$), but a significant main effect of timepoint (*ATS* (1) = 140.64, $p = 1.92E-32$), indicating higher SUDS ratings at the start of scene 1 compared to end of scene 10. These findings suggest that the more pronounced reduction in SUDS ratings from level 1 to level 10 in the VPA group was driven by the significantly higher SUDS ratings in scene 1 timepoint beginning in the VPA group and does not indicate a higher overall within-session exposure in the VPA + retrieval group compared to the placebo + retrieval group.
**HR:** No significant three-way interaction between time point, scene, and group ($p = 0.81$), and no significant two-fold interaction between group and time point ($p = 0.54$), or between group and scene ($p = 0.17$) was detected. However, there was a significant interaction between time point and scene (*ATS* (6.63) = 4.86, $p = 2.69E-05$), indicating different amounts of HR reductions from the beginning to the end across scenes. There was no significant main effect of group ($p = 0.38$) or timepoint ($p = 0.28$), but there was a significant main effect of scene (*ATS* (7.25) = 4.21, $p = 9.34E-05$).
Comparing HR scene 1 to HR scene 10, no statistically significant three-way interaction was observed among timepoint, scene, and group allocation ($p = 0.91$), and there were no significant two-way interactions between group and time point, group and scene, or between time point and scene (all $p > 0.27$). A nominal significant main effect of scene was found (*ATS* (1) = 6.23, $p = 0.013$), while there were no significant main effects of group ($p = 0.058$) or timepoint ($p = 0.14$).
**SCL:** No significant three-fold interaction was detected ($p = 0.95$), nor a significant two-fold interaction between group and scene ($p = 0.97$) or group and time point ($p = 0.55$). However, there was a significant interaction between time point and scene (*WTS* (6.43) = 4.21, $p = 2.05E-04$), indicating different amounts of SCL reductions from the beginning to the end across scenes. No significant main effect of group was present ($p = 0.52$), but there was a significant main effect of scene (*WTS* (6.97) = 15.94, $p = 5.27E-21$) and time point (*WTS* (1) = 41.61, $p = 1.11E-10$).
Comparing SCL scene 1 to SCL scene 10, no statistically significant three-way interaction was observed among timepoint, scene, and group allocation ($p = 0.31$), nor were there any significant two-way interactions between group and timepoint or group and scene or between time point and scene (all $p > 0.29$). There was a significant main effect of time point (*ATS* (1) = 14.46, $p = 0.00014$), but no significant main effect of group ($p = 0.99$) or level ($p = 0.06$).

### Supplementary Tables

ST1. Descriptive statistics of primary and secondary outcomes, effect sizes and p-values.
ST2. Descriptives of the nonparametric ANOVA-type statistic on effect of exposure therapy on primary and secondary outcomes.
ST3. Results of the nonparametric ANOVA-type statistic on effect of exposure therapy on primary and secondary outcomes.
ST4. Adverse events after medication intake at visit 2.
ST5. Complete list of adverse events at visit 2 and visit 3 for the participants of the VPA + retrieval and the placebo + retrieval group.

**Supplementary Table ST1.** Descriptive statistics of primary and secondary outcomes, effect sizes and p-values.

| Outcome | Time | Group | *n* | Mean | Median | *SD* | Min | Max | Cohen's *d* | *p* | Effect size |
|---|---|---|---|---|---|---|---|---|---|---|---|
| **Approach behavior BAT in vivo** | **Baseline** | **Placebo** | 26 | 3.96 | 4.00 | 1.40 | 1.00 | 6.00 | | | |
| | **Baseline** | **VPA** | 22 | 3.77 | 3.50 | 1.31 | 1.00 | 6.00 | -0.14 | | |
| | **Follow-up** | **Placebo** | 26 | 5.23 | 5.00 | 2.12 | 1.00 | 8.00 | | | |
| | **Follow-up** | **VPA** | 22 | 5.41 | 5.00 | 1.53 | 2.00 | 8.00 | 0.10 | | |
| | **Difference score** | **Placebo** | 26 | 1.27 | 1.00 | 1.80 | -2.00 | 6.00 | | | |
| | **Difference score** | **VPA** | 22 | 1.64 | 1.50 | 1.50 | -1.00 | 5.00 | 0.22 | 0.46 | *d* = 0.23 |
| **SUDS after BAT in vivo** | **Baseline** | **Placebo** | 26 | 6.46 | 6.50 | 2.06 | 3.00 | 10.00 | | | |
| | **Baseline** | **VPA** | 22 | 6.86 | 7.00 | 1.93 | 3.00 | 10.00 | 0.20 | | |
| | **Follow-up** | **Placebo** | 26 | 4.08 | 4.00 | 2.61 | 0.00 | 8.00 | | | |
| | **Follow-up** | **VPA** | 22 | 4.55 | 4.50 | 2.18 | 1.00 | 8.00 | 0.19 | | |
| | **Difference score** | **Placebo** | 26 | -2.38 | -2.00 | 2.97 | -9.00 | 3.00 | | | |
| | **Difference score** | **VPA** | 22 | -2.32 | -2.00 | 2.50 | -7.00 | 2.00 | -0.03 | 0.92 | *d* = -0.03 |
| **Approach behavior BAT in VR** | **Baseline** | **Placebo** | 26 | 2.31 | 2.48 | 0.99 | 0.13 | 4.48 | | | |
| | **Baseline** | **VPA** | 22 | 2.49 | 2.24 | 0.69 | 1.35 | 3.81 | 0.20 | | |
| | **Follow-up** | **Placebo** | 26 | 1.42 | 1.46 | 0.99 | 0.00 | 3.69 | | | |
| | **Follow-up** | **VPA** | 22 | 1.44 | 1.53 | 0.69 | 0.00 | 3.03 | 0.03 | | |
| | **Difference score** | **Placebo** | 26 | -0.89 | -0.81 | 0.76 | -3.08 | 0.04 | | | |
| | **Difference score** | **VPA** | 22 | -1.04 | -0.75 | 0.79 | -3.21 | -0.20 | 0.19 | 0.59 | *V* = 0.08 |
| **SUDS BAT in VR** | **Baseline** | **Placebo** | 26 | 7.04 | 7.00 | 2.39 | 1.00 | 10.00 | | | |
| | **Baseline** | **VPA** | 22 | 7.68 | 7.50 | 1.59 | 5.00 | 10.00 | 0.31 | | |
| | **Follow-up** | **Placebo** | 26 | 3.04 | 2.00 | 2.34 | 0.00 | 8.00 | | | |
| | **Follow-up** | **VPA** | 22 | 3.55 | 3.50 | 1.82 | 0.00 | 6.00 | 0.24 | | |
| | **Difference score** | **Placebo** | 26 | -4.00 | -4.00 | 2.56 | -10.00 | 0.00 | | | |
| | **Difference score** | **VPA** | 22 | -4.14 | -4.00 | 2.53 | -10.00 | 1.00 | 0.05 | 0.88 | *d* = 0.05 |
| **FSQ** | **Baseline** | **Placebo** | 26 | 70.62 | 70.50 | 16.74 | 37.00 | 97.00 | | | |
| | **Baseline** | **VPA** | 22 | 72.68 | 71.00 | 14.08 | 45.00 | 99.00 | 0.13 | | |

| | | | | | | | | | | | |
|---|---|---|---|---|---|---|---|---|---|---|---|
| | **Follow-up** | **Placebo** | 26 | 49.54 | 50.00 | 25.53 | 3.00 | 86.00 | | | |
| | **Follow-up** | **VPA** | 22 | 34.09 | 30.00 | 17.97 | 6.00 | 75.00 | -0.69 | | |
| | **Difference score** | **Placebo** | 26 | -21.08 | -14.00 | 21.74 | -72.00 | 20.00 | | | |
| | **Difference score** | **VPA** | 22 | -38.59 | -36.50 | 15.34 | -66.00 | -5.00 | 0.92 | 0.003 | *d* = 0.95 |
| **SBQ** [a] | **Baseline** | **Placebo** | 26 | 54.84 | 59.58 | 19.85 | 4.17 | 90.54 | | | |
| | **Baseline** | **VPA** | 21 | 55.45 | 51.35 | 11.96 | 31.04 | 77.71 | 0.04 | | |
| | **Follow-up** | **Placebo** | 26 | 43.94 | 47.92 | 22.38 | 0.83 | 80.42 | | | |
| | **Follow-up** | **VPA** | 22 | 31.38 | 31.88 | 16.00 | 10.63 | 71.88 | -0.64 | | |
| | **Difference score** | **Placebo** | 26 | -10.89 | -10.64 | 12.75 | -44.63 | 9.17 | | | |
| | **Difference score** | **VPA** | 21 | -24.53 | -22.50 | 15.49 | -54.17 | 0.52 | 0.97 | 0.002 | *d* = 1.02 |
| **DSM-IV impairment** | **Baseline** | **Placebo** | 26 | 5.15 | 5.00 | 1.16 | 3.00 | 7.00 | | | |
| | **Baseline** | **VPA** | 22 | 5.59 | 6.00 | 1.22 | 4.00 | 7.00 | 0.37 | | |
| | **Follow-up** | **Placebo** | 26 | 4.31 | 4.50 | 1.95 | 0.00 | 8.00 | | | |
| | **Follow-up** | **VPA** | 22 | 3.77 | 4.00 | 1.80 | 0.00 | 7.00 | -0.28 | | |
| | **Difference score** | **Placebo** | 26 | -0.85 | -1.00 | 1.78 | -4.00 | 2.00 | | | |
| | **Difference score** | **VPA** | 22 | -1.82 | -1.50 | 1.94 | -6.00 | 1.00 | 0.52 | 0.07 | *d* = 0.56 |
| **DSM-IV distress** | **Baseline** | **Placebo** | 26 | 5.38 | 6.00 | 1.27 | 2.00 | 7.00 | | | |
| | **Baseline** | **VPA** | 22 | 4.77 | 5.00 | 1.15 | 2.00 | 7.00 | -0.50 | | |
| | **Follow-up** | **Placebo** | 26 | 3.50 | 4.00 | 1.79 | 0.00 | 6.00 | | | |
| | **Follow-up** | **VPA** | 22 | 3.18 | 3.00 | 1.53 | 0.00 | 6.00 | -0.19 | | |
| | **Difference score** | **Placebo** | 26 | -1.88 | -2.00 | 1.80 | -5.00 | 1.00 | | | |
| | **Difference score** | **VPA** | 22 | -1.59 | -2.00 | 1.50 | -4.00 | 1.00 | -0.18 | 0.52 | *d* = -0.20 |
| **STAI-S** | **Baseline** | **Placebo** | 26 | 33.58 | 31.50 | 7.70 | 22.00 | 54.00 | | | |
| | **Baseline** | **VPA** | 22 | 32.41 | 30.50 | 8.03 | 22.00 | 57.00 | -0.15 | | |
| | **Follow-up** | **Placebo** | 26 | 33.92 | 33.50 | 8.64 | 20.00 | 54.00 | | | |
| | **Follow-up** | **VPA** | 22 | 30.50 | 29.00 | 6.20 | 22.00 | 50.00 | -0.45 | | |
| | **Difference score** | **Placebo** | 26 | 0.35 | 1.00 | 6.51 | -10.00 | 15.00 | | | |
| | **Difference score** | **VPA** | 22 | -1.91 | -2.50 | 5.88 | -18.00 | 10.00 | -0.36 | 0.23 | *d* = -0.37 |
| **HR BAT in VR** [b] | **Baseline** | **Placebo** | 22 | 1.77 | 0.14 | 8.05 | 1.72 | -10.30 | | | |

| | | | | | | | | | | | |
|---|---|---|---|---|---|---|---|---|---|---|---|
| | **Baseline** | **VPA** | 17 | 3.44 | 3.09 | 6.41 | 1.56 | -5.88 | -0.23 | | |
| | **Follow-up** | **Placebo** | 21 | -0.34 | -0.34 | 4.37 | 0.95 | -9.83 | | | |
| | **Follow-up** | **VPA** | 19 | -0.36 | 0.13 | 2.87 | 0.66 | -6.11 | 0.00 | | |
| | **Difference score** | **Placebo** | 18 | -3.80 | -3.33 | 8.54 | -18.69 | 12.15 | | | |
| | **Difference score** | **VPA** | 14 | -3.79 | -4.73 | 7.73 | -21.38 | 11.87 | -0.00 | 0.84 | *d* = -0.08 |
| **SCL BAT in VR** [b] | **Baseline** | **Placebo** | 26 | 1.03 | 0.28 | 2.86 | 0.56 | -0.92 | | | |
| | **Baseline** | **VPA** | 21 | 0.35 | 0.20 | 0.47 | 0.10 | -0.03 | -0.32 | | |
| | **Follow-up** | **Placebo** | 25 | 0.04 | 0.00 | 0.40 | 0.08 | -1.07 | | | |
| | **Follow-up** | **VPA** | 26 | 1.03 | 0.28 | 2.86 | 0.56 | -0.92 | 0.16 | | |
| | **Difference score** | **Placebo** | 25 | -1.02 | -0.13 | 3.00 | -14.41 | 0.86 | | | |
| | **Difference score** | **VPA** | 21 | -0.26 | -0.09 | 0.59 | -2.10 | 0.62 | -0.34 | 0.019 | *V* = 0.35 |
| **Anxiety rating of spider pictures** [c, d] | **Baseline** | **Placebo** | 22 | 78.06 | 81.53 | 16.96 | 28.63 | 100.00 | | | |
| | **Baseline** | **VPA** | 19 | 77.20 | 82.38 | 19.19 | 34.81 | 100.00 | 0.05 | | |
| | **Follow-up** | **Placebo** | 22 | 62.05 | 64.56 | 28.12 | 3.19 | 93.13 | | | |
| | **Follow-up** | **VPA** | 19 | 55.96 | 64.00 | 30.18 | 0.00 | 95.44 | -0.21 | | |
| | **Difference score** | **Placebo** | 22 | -16.01 | -8.91 | 23.39 | -68.38 | 14.19 | | | |
| | **Difference score** | **VPA** | 19 | -21.23 | -10.75 | 25.08 | -79.38 | 9.19 | 0.22 | 0.50 | *d* = 0.22 |
| **Valence rating of spider pictures** [c, d] | **Baseline** | **Placebo** | 22 | 86.87 | 89.19 | 12.86 | 47.69 | 100.00 | | | |
| | **Baseline** | **VPA** | 19 | 87.04 | 89.56 | 10.19 | 66.44 | 100.00 | 0.01 | | |
| | **Follow-up** | **Placebo** | 22 | 75.89 | 86.75 | 26.62 | 9.44 | 99.94 | | | |
| | **Follow-up** | **VPA** | 19 | 76.88 | 82.19 | 19.80 | 36.81 | 100.00 | 0.04 | | |
| | **Difference score** | **Placebo** | 22 | -10.97 | -5.84 | 19.92 | -58.38 | 17.19 | | | |
| | **Difference score** | **VPA** | 19 | -10.15 | -3.81 | 19.10 | -57.81 | 12.94 | -0.04 | 0.84 | *d* = -0.07 |
| **Arousal rating of spider pictures** [c, d] | **Baseline** | **Placebo** | 22 | 78.65 | 79.88 | 17.23 | 21.13 | 100.00 | | | |
| | **Baseline** | **VPA** | 19 | 79.89 | 84.81 | 14.86 | 47.06 | 99.94 | 0.08 | | |
| | **Follow-up** | **Placebo** | 22 | 66.80 | 69.31 | 26.54 | 1.81 | 96.13 | | | |
| | **Follow-up** | **VPA** | 19 | 61.52 | 68.56 | 26.09 | 3.25 | 97.38 | -0.20 | | |
| | **Difference score** | **Placebo** | 22 | -11.85 | -9.09 | 20.77 | -68.56 | 16.63 | | | |
| | **Difference score** | **VPA** | 19 | -18.37 | -11.06 | 25.62 | -85.00 | 15.38 | 0.28 | 0.80 | *V* = 0.04 |

| | | | | | | | | | | | |
|---|---|---|---|---|---|---|---|---|---|---|---|
| **SCL spider pictures** [b-d] | **Baseline** | **Placebo** | 21 | 0.15 | 0.05 | 0.21 | 0.05 | 0.00 | | | |
| | **Baseline** | **VPA** | 18 | 0.14 | 0.02 | 0.43 | 0.10 | -0.09 | -0.01 | | |
| | **Follow-up** | **Placebo** | 22 | 0.11 | 0.03 | 0.22 | 0.05 | -0.16 | | | |
| | **Follow-up** | **VPA** | 20 | 0.04 | 0.01 | 0.13 | 0.03 | -0.04 | -0.35 | | |
| | **Difference score** | **Placebo** | 20 | -0.04 | -0.01 | 0.32 | -0.64 | 0.78 | | | |
| | **Difference score** | **VPA** | 18 | -0.09 | -0.01 | 0.29 | -1.23 | 0.05 | 0.18 | 0.46 | $V = 0.12$ |
| **HR spider pictures** [b-d] | **Baseline** | **Placebo** | 18 | 0.69 | 0.27 | 2.95 | -4.66 | 7.61 | | | |
| | **Baseline** | **VPA** | 17 | -0.86 | -0.75 | 3.17 | -6.83 | 4.02 | 0.50 | | |
| | **Follow-up** | **Placebo** | 20 | -0.73 | -1.58 | 2.77 | -4.75 | 4.98 | | | |
| | **Follow-up** | **VPA** | 19 | -2.10 | -2.30 | 2.54 | -8.60 | 2.06 | 0.51 | | |
| | **Difference score** | **Placebo** | 17 | -1.96 | -1.68 | 2.77 | -7.63 | 1.47 | | | |
| | **Difference score** | **VPA** | 16 | -1.56 | -1.40 | 3.45 | -7.56 | 4.36 | 0.13 | 0.06 | $V = 0.33$ |
| **Startle spider pictures** [b-d] | **Baseline** | **Placebo** | 22 | 59.69 | 47.80 | 45.30 | 9.66 | 8.39 | | | |
| | **Baseline** | **VPA** | 18 | 58.63 | 34.46 | 51.15 | 12.06 | 17.86 | 0.02 | | |
| | **Follow-up** | **Placebo** | 21 | 50.69 | 37.39 | 42.58 | 9.29 | 1.22 | | | |
| | **Follow-up** | **VPA** | 19 | 36.80 | 22.56 | 40.67 | 9.33 | 0.86 | 0.33 | | |
| | **Difference score** | **Placebo** | 20 | -11.52 | -13.88 | 33.43 | -61.85 | 83.04 | | | |
| | **Difference score** | **VPA** | 18 | -20.56 | -11.87 | 46.38 | -127.35 | 79.20 | -0.23 | 0.77 | $V = 0.05$ |
| **Level of VPA in blood at visit 2** [a] | **before study medication** | **Placebo** | 24 | 0.00 | 0.00 | 0.00 | 0.00 | 0.00 | | | |
| | **before study medication** | **VPA** | 20 | 0.00 | 0.00 | 0.00 | 0.00 | 0.00 | 0.00 | | |
| | **end of visit 2** | **Placebo** | 24 | 0.00 | 0.00 | 0.00 | 0.00 | 0.00 | | | |
| | **end of visit 2** | **VPA** | 20 | 40.27 | 38.75 | 6.28 | 25.20 | 49.60 | -9.54 | < 2.2e-16 | |

VPA = valproic acid, FSQ = Fear of Spiders Questionnaire, SBQ = Spider Phobia Beliefs Questionnaire, BAT = behavioral approach test, STAI-S = State Trait Anxiety Inventory – State, HR = heart rate, SCL = skin conductance level. Cohen's *d* refers to the effect size calculated based on mean values. *p* value refers to *p* value of the main effect group allocation of the used statistical model, i.e. difference score models (follow-up, baseline). These models were either calculated with linear models, or if assumptions were violated, then dependent variables were revisualized for the covariates age and sex. Subsequently, nonparametric Mann–Whitney tests for independent samples were applied to test for group differences. Effect size refers to the effect size of the main effect group allocation of the used statistical model, i.e. statistical model with age and sex as covariates included.

In the table only participants with data points at baseline and follow-up are included, reasons for missing data points are the following:
[a] not collected at baseline
[b] corrupted recordings of physiological data due to temporary ECG-electrode dysfunction or excessive movement of participants on individual test days
[c] technical problems with the conduction of PPST on individual test days (VPA: $n = 1$, placebo: $n = 1$)
[d] participants diagnosed with an additional specific phobia (animal type: snakes) as pictures of snakes were included as a control condition in the PPST (VPA: $n = 2$, placebo: $n = 3$)

**Supplementary Table ST2.** Descriptives of the nonparametric ANOVA-type statistic on effect of exposure therapy on primary and secondary outcomes.

| | Group | time | Nobs | RankMeans | RTE |
|---|---|---|---|---|---|
| **Approach behavior BAT in vivo.** | **Placebo** | **Baseline** | 26 | 36.31 | 0.37 |
| | **Placebo** | **Follow-up** | 26 | 60.58 | 0.63 |
| | **VPA** | **Baseline** | 22 | 31.34 | 0.32 |
| | **VPA** | **Follow-up** | 22 | 65.80 | 0.68 |
| **SUDS after BAT in vivo** | **Placebo** | **Baseline** | 26 | 59.04 | 0.61 |
| | **Placebo** | **Follow-up** | 26 | 33.79 | 0.35 |
| | **VPA** | **Baseline** | 22 | 64.00 | 0.66 |
| | **VPA** | **Follow-up** | 22 | 37.93 | 0.39 |
| **Anxiety rating of spider pictures** [c, d] | **Placebo** | **Baseline** | 22 | 53.23 | 0.64 |
| | **Placebo** | **Follow-up** | 22 | 32.18 | 0.39 |
| | **VPA** | **Baseline** | 19 | 53.47 | 0.65 |
| | **VPA** | **Follow-up** | 19 | 26.74 | 0.32 |
| **Valence rating of spider pictures** [c, d] | **Placebo** | **Baseline** | 22 | 50.73 | 0.61 |
| | **Placebo** | **Follow-up** | 22 | 33.95 | 0.41 |
| | **VPA** | **Baseline** | 19 | 48.87 | 0.59 |
| | **VPA** | **Follow-up** | 19 | 32.18 | 0.39 |
| **Arousal rating of spider pictures** [c, d] | **Placebo** | **Baseline** | 22 | 50.45 | 0.61 |
| | **Placebo** | **Follow-up** | 22 | 34.18 | 0.41 |
| | **VPA** | **Baseline** | 19 | 52.84 | 0.64 |
| | **VPA** | **Follow-up** | 19 | 28.26 | 0.34 |
| **SCL spider pictures** [b-d] | **Placebo** | **Baseline** | 20 | 44.45 | 0.58 |
| | **Placebo** | **Follow-up** | 20 | 36.00 | 0.47 |
| | **VPA** | **Baseline** | 18 | 43.28 | 0.56 |
| | **VPA** | **Follow-up** | 18 | 29.89 | 0.39 |
| **HR spider pictures** [b-d] | **Placebo** | **Baseline** | 17 | 44.29 | 0.66 |
| | **Placebo** | **Follow-up** | 17 | 26.88 | 0.40 |

| | | | | | |
|---|---|---|---|---|---|
| | **VPA** | **Baseline** | 16 | 39.06 | 0.58 |
| | **VPA** | **Follow-up** | 16 | 23.50 | 0.35 |
| **Startle spider pictures** [b-d] | **Placebo** | **Baseline** | 20 | 48.15 | 0.63 |
| | **Placebo** | **Follow-up** | 20 | 32.50 | 0.42 |
| | **VPA** | **Baseline** | 18 | 46.39 | 0.60 |
| | **VPA** | **Follow-up** | 18 | 26.56 | 0.34 |
| **FSQ** | **Placebo** | **Baseline** | 26 | 63.33 | 0.65 |
| | **Placebo** | **Follow-up** | 26 | 40.46 | 0.42 |
| | **VPA** | **Baseline** | 22 | 65.77 | 0.68 |
| | **VPA** | **Follow-up** | 22 | 23.20 | 0.24 |
| **SBQ** [a] | **Placebo** | **Baseline** | 26 | 61.69 | 0.65 |
| | **Placebo** | **Follow-up** | 26 | 37.88 | 0.40 |
| | **VPA** | **Baseline** | 21 | 68.81 | 0.73 |
| | **VPA** | **Follow-up** | 21 | 20.52 | 0.21 |
| **DSM-IV impairment** | **Placebo** | **Baseline** | 26 | 55.73 | 0.58 |
| | **Placebo** | **Follow-up** | 26 | 41.35 | 0.43 |
| | **VPA** | **Baseline** | 22 | 65.09 | 0.67 |
| | **VPA** | **Follow-up** | 22 | 31.82 | 0.33 |
| **DSM-IV distress** | **Placebo** | **Baseline** | 26 | 69.65 | 0.72 |
| | **Placebo** | **Follow-up** | 26 | 34.50 | 0.35 |
| | **VPA** | **Baseline** | 22 | 58.95 | 0.61 |
| | **VPA** | **Follow-up** | 22 | 29.59 | 0.30 |
| **STAI-S** | **Placebo** | **Baseline** | 26 | 49.85 | 0.51 |
| | **Placebo** | **Follow-up** | 26 | 52.38 | 0.54 |
| | **VPA** | **Baseline** | 22 | 53.66 | 0.55 |
| | **VPA** | **Follow-up** | 22 | 37.16 | 0.38 |
| **SUDS BAT in VR** | **Placebo** | **Baseline** | 26 | 65.87 | 0.68 |
| | **Placebo** | **Follow-up** | 26 | 27.27 | 0.28 |

| | | | | | |
|---|---|---|---|---|---|
| | **VPA** | **Baseline** | 22 | 71.36 | 0.74 |
| | **VPA** | **Follow-up** | 22 | 30.20 | 0.31 |
| **Approach behavior BAT in VR** | **Placebo** | **Baseline** | 26 | 63.81 | 0.66 |
| | **Placebo** | **Follow-up** | 26 | 31.62 | 0.32 |
| | **VPA** | **Baseline** | 22 | 68.95 | 0.71 |
| | **VPA** | **Follow-up** | 22 | 29.91 | 0.31 |
| **HR BAT in VR** [b] | **Placebo** | **Baseline** | 18 | 36.22 | 0.56 |
| | **Placebo** | **Follow-up** | 18 | 28.06 | 0.43 |
| | **VPA** | **Baseline** | 14 | 38.71 | 0.60 |
| | **VPA** | **Follow-up** | 14 | 27.21 | 0.42 |
| **SCL BAT in VR** [b] | **Placebo** | **Baseline** | 25 | 56.28 | 0.61 |
| | **Placebo** | **Follow-up** | 25 | 32.28 | 0.35 |
| | **VPA** | **Baseline** | 21 | 58.05 | 0.63 |
| | **VPA** | **Follow-up** | 21 | 40.24 | 0.43 |

VPA = valproic acid, FSQ = Fear of Spiders Questionnaire, SBQ = Spider Phobia Beliefs Questionnaire, BAT = behavioral approach test, STAI-S = State Trait Anxiety Inventory – State, HR = heart rate, SCL = skin conductance level.

In the table, only participants with data points at baseline and follow-up are included; reasons for missing data points are the following:
[a] not collected at baseline
[b] technical problems with the collection of physiological data on individual test days
[c] technical problems with the conduction of PPST on individual test days (VPA: $n$ = 1, placebo: $n$ = 1)
[d] participants diagnosed with an additional specific phobia (animal type: snakes) as pictures of snakes were included as a control condition in the PPST (VPA: $n$ = 2, placebo: $n$ = 3)

**Supplementary Table ST3.** Results of the nonparametric ANOVA-type statistic on effect of exposure therapy on primary and secondary outcomes.

| | Df | *F* | *p* |
|---|---|---|---|
| ***Approach behavior BAT in vivo*** | | | |
| **Group** | 1 | 0.00 | 0.98 |
| **Visit** | 1 | 40.73 | 1.75E-10 |
| **Group x Visit** | 1 | 1.23 | 0.27 |
| ***SUDS after BAT in vivo*** | | | |
| **Group** | 1 | 0.65 | 0.42 |
| **Visit** | 1 | 33.46 | 7.27E-09 |
| **Group x Visit** | 1 | 0.01 | 0.93 |
| ***Anxiety rating of spider pictures*** | | | |
| **Group** | 1 | 0.32 | 0.57 |
| **Visit** | 1 | 26.65 | 2.44E-07 |
| **Group x Visit** | 1 | 0.38 | 0.54 |
| ***Valence rating of spider pictures*** | | | |
| **Group** | 1 | 0.18 | 0.67 |
| **Visit** | 1 | 8.71 | 3.17E-03 |
| **Group x Visit** | 1 | 0.00 | 0.99 |
| ***Arousal rating spider pictures*** | | | |
| **Group** | 1 | 0.13 | 0.72 |
| **Visit** | 1 | 18.34 | 1.85E-05 |
| **Group x Visit** | 1 | 0.76 | 0.38 |
| ***SCL spider pictures*** | | | |
| **Group** | 1 | 1.67 | 0.20 |
| **Visit** | 1 | 2.96 | 0.09 |
| **Group x Visit** | 1 | 0.15 | 0.70 |
| ***HR spider pictures*** | | | |
| **Group** | 1 | 1.33 | 0.25 |
| **Visit** | 1 | 11.45 | 7.16E-04 |
| **Group x Visit** | 1 | 0.04 | 0.85 |
| ***Startle spider pictures*** | | | |
| **Group** | 1 | 1.17 | 0.28 |
| **Visit** | 1 | 9.82 | 1.73E-03 |
| **Group x Visit** | 1 | 0.14 | 0.71 |
| ***FSQ*** | | | |
| **Group** | 1 | 1.84 | 0.17 |
| **Visit** | 1 | 108.49 | 2.09E-25 |
| **Group x Visit** | 1 | 9.84 | 1.71E-03 |

| | | | |
|---|---|---|---|
| | ***post-hoc tests for each group separately*** | | |
| **within VPA group: visit** | 1 | 132.19 | 1.36E-30 |
| **within placebo group: visit** | 1 | 24.03 | 9.47E-07 |
| ***SBQ*** | | | |
| **Group** | 1 | 1.45 | 0.23 |
| **Visit** | 1 | 85.97 | 1.82E-20 |
| **Group x Visit** | 1 | 9.91 | 1.64E-03 |
| | ***post-hoc tests for each group separately*** | | |
| **within VPA group: visit** | 1 | 59.42 | 1.27E-14 |
| **within placebo group: visit** | 1 | 23.17 | 1.49E-06 |
| ***DSM-IV impairment*** | | | |
| **Group** | 1 | 0.00 | 0.99 |
| **Visit** | 1 | 21.39 | 3.74E-06 |
| **Group x Visit** | 1 | 3.36 | 0.07 |
| ***DSM-IV distress*** | | | |
| **Group** | 1 | 2.64 | 0.10 |
| **Visit** | 1 | 56.10 | 6.87E-14 |
| **Group x Visit** | 1 | 0.45 | 0.50 |
| ***STAI-S*** | | | |
| **Group** | 1 | 3.30 | 0.07 |
| **Visit** | 1 | 0.94 | 0.33 |
| **Group x Visit** | 1 | 1.75 | 0.19 |
| ***SUDS BAT in VR*** | | | |
| **Group** | 1 | 0.93 | 0.33 |
| **Visit** | 1 | 141.97 | 9.89E-33 |
| **Group x Visit** | 1 | 0.15 | 0.70 |
| ***Approach behavior BAT in VR*** | | | |
| **Group** | 1 | 0.12 | 0.73 |
| **Visit** | 1 | 94.88 | 2.02E-22 |
| **Group x Visit** | 1 | 0.88 | 0.35 |
| ***HR BAT in VR*** | | | |
| **Group** | 1 | 0.04 | 0.85 |
| **Visit** | 1 | 4.34 | 0.04 |
| **Group x Visit** | 1 | 0.12 | 0.72 |
| ***SCL BAT in VR*** | | | |
| **Group** | 1 | 1.18 | 0.28 |
| **Visit** | 1 | 13.78 | 2.06E-04 |
| **Group x Visit** | 1 | 0.30 | 0.58 |

**Supplementary Table ST4.** Adverse events after medication intake at visit 2.

| | **Placebo** | **VPA** |
|---|---|---|
| **Gastrointestinal System** | | |
| Nausea | | 1 |
| **Central and Peripheral Nervous System** | | |
| Headache | 2 | |
| Dizziness | 1 | |
| Drowsiness | 2 | |
| Tiredness | 8 | 7 |
| **Cardiovascular** | | |
| Hypertension | 6 | 4 |
| Hypotension | 1 | 1 |
| Circulatory complaints | | 1 |
| **Overall body** | | |
| Skin rash | 1 | |
| Muscle fatigue | 2 | 1 |
| **Sum total** | **23** | **15** |

**Supplementary Table ST5.** Complete list of adverse events assessed at visit 2 and visit 3 for the participants of the VPA + retrieval and the placebo + retrieval group.

| Participant | Adverse event | Extent | Relatedness | Visit |
|---|---|---|---|---|
| 100 | Hypertension | Mild | Possible | Intervention |
| 100 | Hypertension | Mild | No | Follow-up |
| 101 | Tiredness | Mild | Possible | Intervention |
| 101 | Hypertension | Mild | No | Follow-up |
| 102 | Tiredness | Mild | No | Intervention |
| 102 | Tiredness | Mild | No | Follow-up |
| 103 | Tiredness | Mild | Possible | Intervention |
| 104 | Hypertension | Mild | Possible | Intervention |
| 104 | Tiredness | Moderate | Possible | Intervention |
| 104 | Headache with Paresthesia | Moderate | Unlikely | Between intervention and follow-up |
| 105 | Tiredness | Mild | No | Intervention |
| 106 | Hypotension | Mild | Unlikely | Intervention |
| 106 | Tiredness | Mild | Unlikely | Intervention |
| 107 | Hypertension | Mild | Possible | Intervention |
| 107 | Hypertension | Mild | No | Follow-up |
| 108 | Hypotension | Mild | Possible | Intervention |
| 108 | Tiredness | Mild | No | Follow-up |
| 108 | Hypotension | Mild | No | Follow-up |
| 109 | Tiredness | Mild | No | Intervention |
| 109 | Hypertension | Mild | No | Intervention |
| 110 | Tiredness | Mild | No | Follow-up |
| 110 | Headache | Mild | No | Follow-up |
| 111 | Tiredness | Mild | No | Intervention |
| 112 | Hypertension | Mild | Unlikely | Intervention |
| 112 | Tiredness | Mild | No | Intervention |
| 112 | Hypertension | Mild | No | Intervention |
| 112 | Hypertension | Mild | No | Follow-up |
| 113 | Hypertension | Mild | Possible | Intervention |
| 113 | Tiredness | Mild | No | Follow-up |
| 114 | Muscle fatigue | Mild | No | Follow-up |
| 114 | Hypertension | Mild | No | Follow-up |
| 115 | Tiredness | Mild | No | Follow-up |
| 115 | Tiredness | Mild | No | Follow-up |
| 116 | Tiredness | Mild | No | Follow-up |
| 116 | Drowsiness | Mild | No | Follow-up |
| 116 | Muscle fatigue | Mild | No | Follow-up |
| 116 | Dizziness | Mild | No | Follow-up |
| 117 | Hypertension | Mild | Unlikely | Intervention |
| 117 | Tiredness | Mild | No | Intervention |
| 117 | Hypertension | Mild | No | Follow-up |

| | | | | |
|---|---|---|---|---|
| 118 | Tiredness | Mild | No | Follow-up |
| 119 | Tiredness | Mild | No | Follow-up |
| 120 | Hypertension | Mild | No | Intervention |
| 120 | Hypertension | Mild | Possible | Intervention |
| 120 | Headache | Mild | Possible | Intervention |
| 120 | Drowsiness | Mild | No | Intervention |
| 120 | Skin rash right arm | Mild | Possible | Intervention |
| 120 | Tiredness | Mild | No | Follow-up |
| 130 | Tiredness | Mild | Possible | Intervention |
| 130 | Tiredness | Mild | No | Follow-up |
| 130 | Hypertension | Mild | No | Follow-up |
| 131 | Hypertension | Mild | No | Intervention |
| 131 | Headache | Mild | Possible | Intervention |
| 131 | Drowsiness | Mild | Possible | Intervention |
| 131 | Tiredness | Mild | Possible | Intervention |
| 131 | Tiredness | Mild | No | Follow-up |
| 132 | Tiredness | Mild | No | Intervention |
| 132 | Tiredness | Mild | No | Follow-up |
| 133 | Hypertension | Mild | Unlikely | Intervention |
| 134 | Tiredness | Mild | No | Intervention |
| 135 | Tiredness | Mild | Possible | Intervention |
| 136 | Tiredness | Mild | No | Follow-up |
| 136 | Muscle fatigue | Mild | No | Follow-up |
| 137 | Tiredness | Mild | No | Follow-up |
| 138 | Tiredness | Mild | Possible | Intervention |
| 138 | Hypotension | Mild | No | Follow-up |
| 139 | Nausea | Mild | Possible | Intervention |
| 139 | Circulatory complaints | Mild | Possible | Intervention |
| 140 | Tiredness | Mild | Possible | Intervention |
| 140 | Dizziness | Mild | Possible | Intervention |
| 140 | Headache | Mild | No | Follow-up |
| 140 | Tiredness | Mild | No | Follow-up |
| 140 | Drowsiness | Mild | No | Follow-up |
| 141 | Tiredness | Mild | No | Follow-up |
| 142 | Hypertension | Mild | Possible | Intervention |
| 142 | Muscle fatigue | Mild | No | Follow-up |
| 143 | Tiredness | Mild | Possible | Intervention |
| 143 | Tiredness | Mild | No | Follow-up |
| 144 | Tiredness | Moderate | No | Follow-up |
| 144 | Drowsiness | Mild | No | Follow-up |
| 145 | Tiredness | Mild | Possible | Intervention |
| 146 | Tiredness | Mild | Possible | Intervention |
| 146 | Drowsiness | Mild | Possible | Intervention |
| 146 | Muscle fatigue | Mild | Possible | Intervention |

| | | | | |
|---|---|---|---|---|
| 146 | Headache | Mild | No | Follow-up |
| 146 | Tiredness | Mild | No | Follow-up |
| 146 | Drowsiness | Mild | No | Follow-up |
| 147 | Tiredness | Mild | Possible | Intervention |
| 147 | Muscle fatigue | Mild | Possible | Intervention |
| 148 | Tiredness | Mild | No | Follow-up |
| 148 | Dizziness | Mild | No | Follow-up |
| 148 | Muscle fatigue | Mild | No | Follow-up |
| 148 | Drowsiness | Mild | No | Follow-up |
| 149 | Hypertension | Mild | Unlikely | Intervention |
| 149 | Tiredness | Mild | Possible | Intervention |
| 149 | Muscle fatigue | Mild | Possible | Intervention |
| 150 | Tiredness | Mild | Possible | Intervention |
| 150 | Tiredness | Mild | No | Follow-up |
| 150 | Skin rash | Mild | Unlikely | Between intervention and follow-up |

Participants from the VPA + retrieval group are marked in gray. *Note*: Participant identifiers listed in this table have been replaced and do not correspond to the actual participant numbers used in the study.

### Supplementary Figures

SF1. Additional secondary outcomes at visit 1 (baseline) and visit 3 (follow-up).
SF2. Scenes 1–10 from the exposure therapy in VR.
SF3. Possible adverse events on visit 2 assessed by visual analogue scales (VAS).
SF4. Subjective Units of Distress Scale (SUDS) (A), skin conductance level (SCL) (B), and heart rate (HR) (C) during exposure therapy in VR.

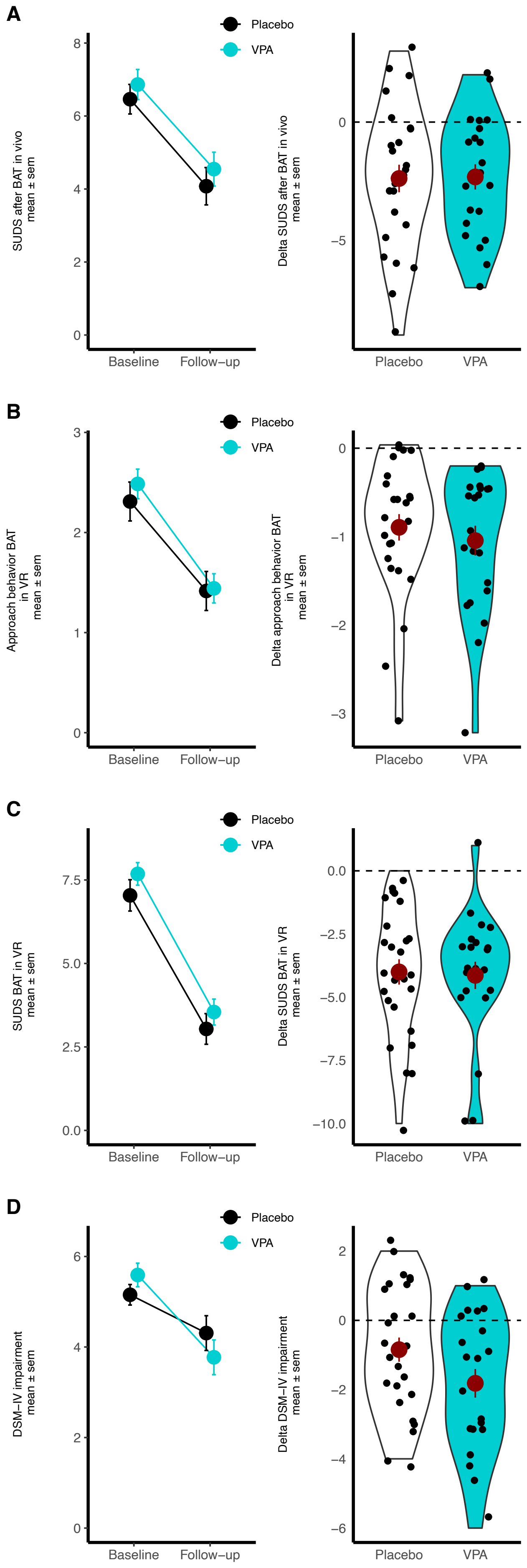
A
B
C
D
Placebo
VPA
SUDS after BAT in vivo
mean ± sem
Delta SUDS after BAT in vivo
mean ± sem
Approach behavior BAT
in VR
mean ± sem
Delta approach behavior BAT
in VR
mean ± sem
SUDS BAT in VR
mean ± sem
Delta SUDS BAT in VR
mean ± sem
DSM−IV impairment
mean ± sem
Delta DSM−IV impairment
mean ± sem
Baseline
Follow−up

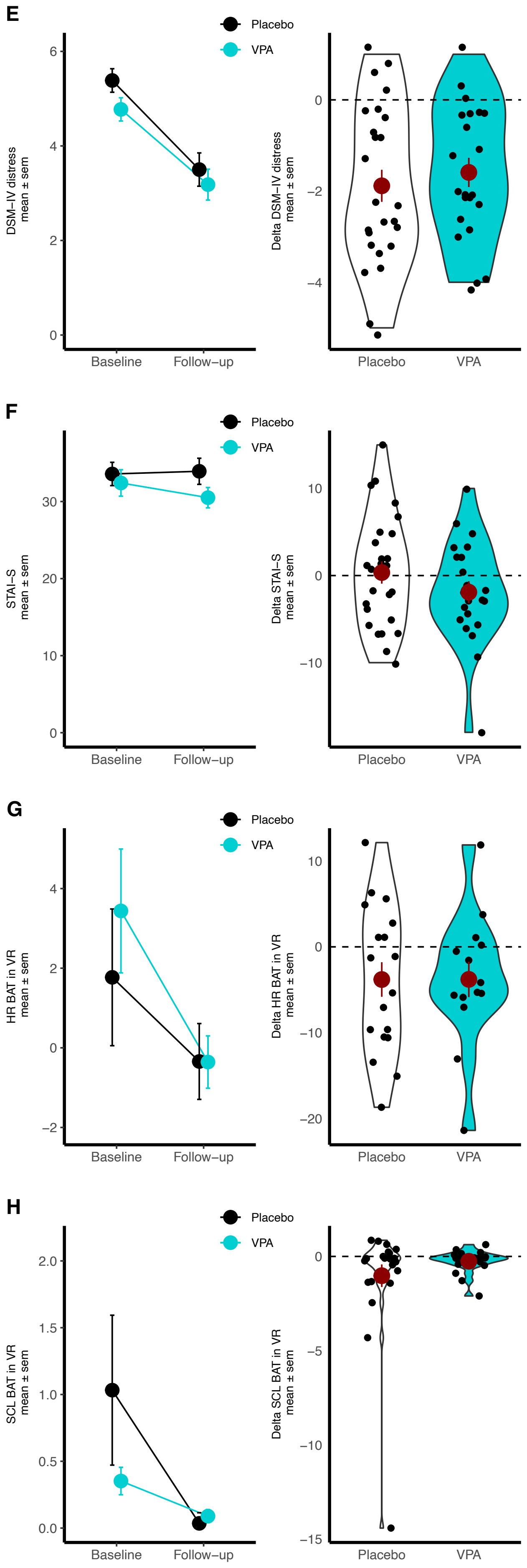
E
Placebo
VPA
DSM–IV distress
mean ± sem
Delta DSM–IV distress
mean ± sem
Baseline
Follow–up
Placebo
VPA
F
STAI–S
mean ± sem
Delta STAI–S
mean ± sem
G
HR BAT in VR
mean ± sem
Delta HR BAT in VR
mean ± sem
H
SCL BAT in VR
mean ± sem
Delta SCL BAT in VR
mean ± sem

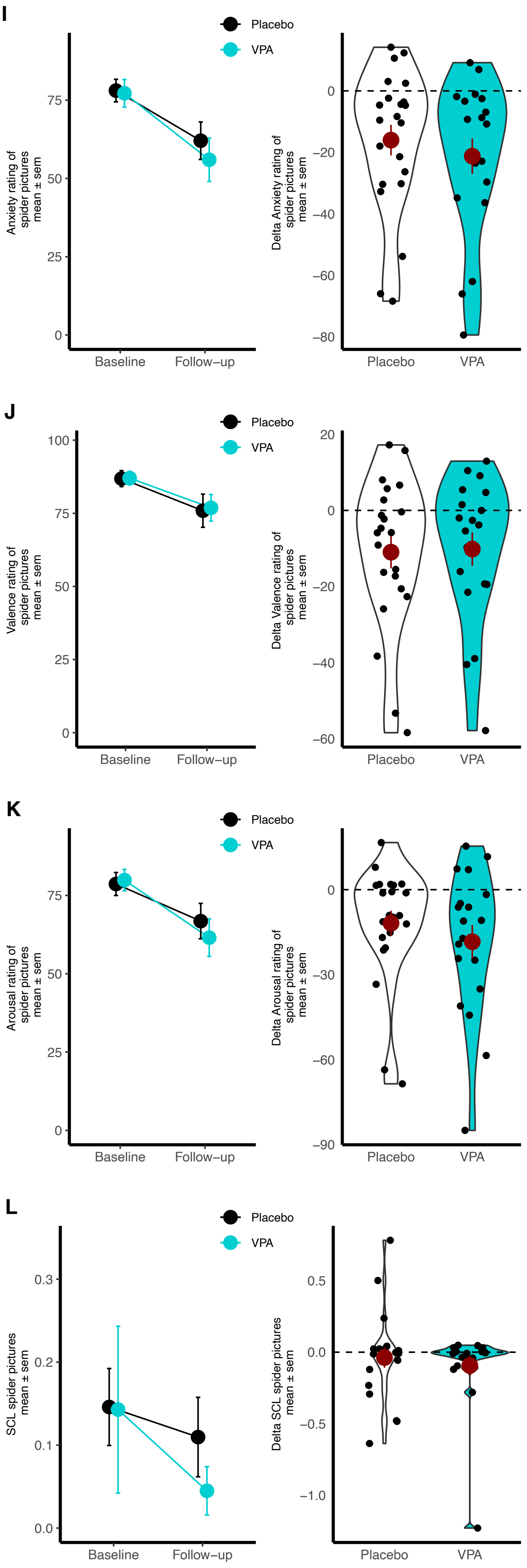
I
Placebo
VPA
Anxiety rating of spider pictures mean ± sem
Baseline
Follow−up
Delta Anxiety rating of spider pictures mean ± sem
Placebo
VPA
J
Valence rating of spider pictures mean ± sem
Delta Valence rating of spider pictures mean ± sem
K
Arousal rating of spider pictures mean ± sem
Delta Arousal rating of spider pictures mean ± sem
L
SCL spider pictures mean ± sem
Delta SCL spider pictures mean ± sem

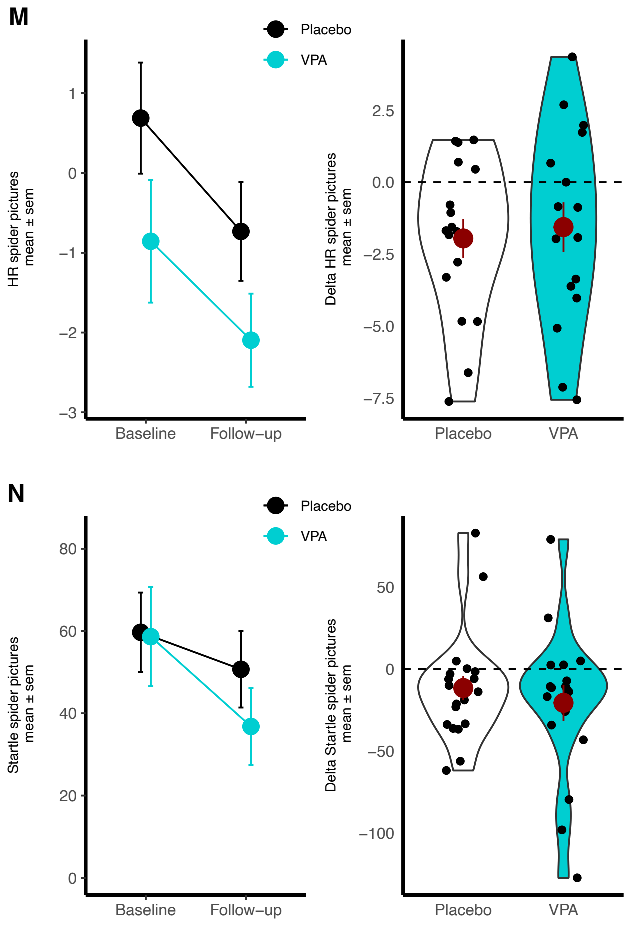
M
Placebo
VPA
HR spider pictures
mean ± sem
1
0
−1
−2
−3
Baseline
Follow−up
Delta HR spider pictures
mean ± sem
2.5
0.0
−2.5
−5.0
−7.5
Placebo
VPA
N
Placebo
VPA
Startle spider pictures
mean ± sem
80
60
40
20
0
Baseline
Follow−up
Delta Startle spider pictures
mean ± sem
50
0
−50
−100
Placebo
VPA

**Supplementary Figure SF1. Additional secondary outcomes at visit 1 (baseline) and visit 3 (follow-up).** The *x* axis represents the two assessment timepoints (baseline, follow-up), while the *y* axis depicts (A) the SUDS after BAT in vivo, (B) approach behavior in VR, (C) the SUDS during BAT in VR, (D) DSM-IV impairment, (E) DSM-IV distress, (F) STAI-S, (G) HR during BAT in VR, (H) SCL during BAT in VR, (I) the anxiety rating of spider pictures, (J) the valence rating of spider pictures, (K) the arousal rating of spider pictures, (L) SCL spider pictures, (M) HR spider pictures, (N) startle spider pictures. The solid line represents the placebo + retrieval group, and the dashed line represents the valproic acid + retrieval group. Displayed are means and the standard errors of the means.

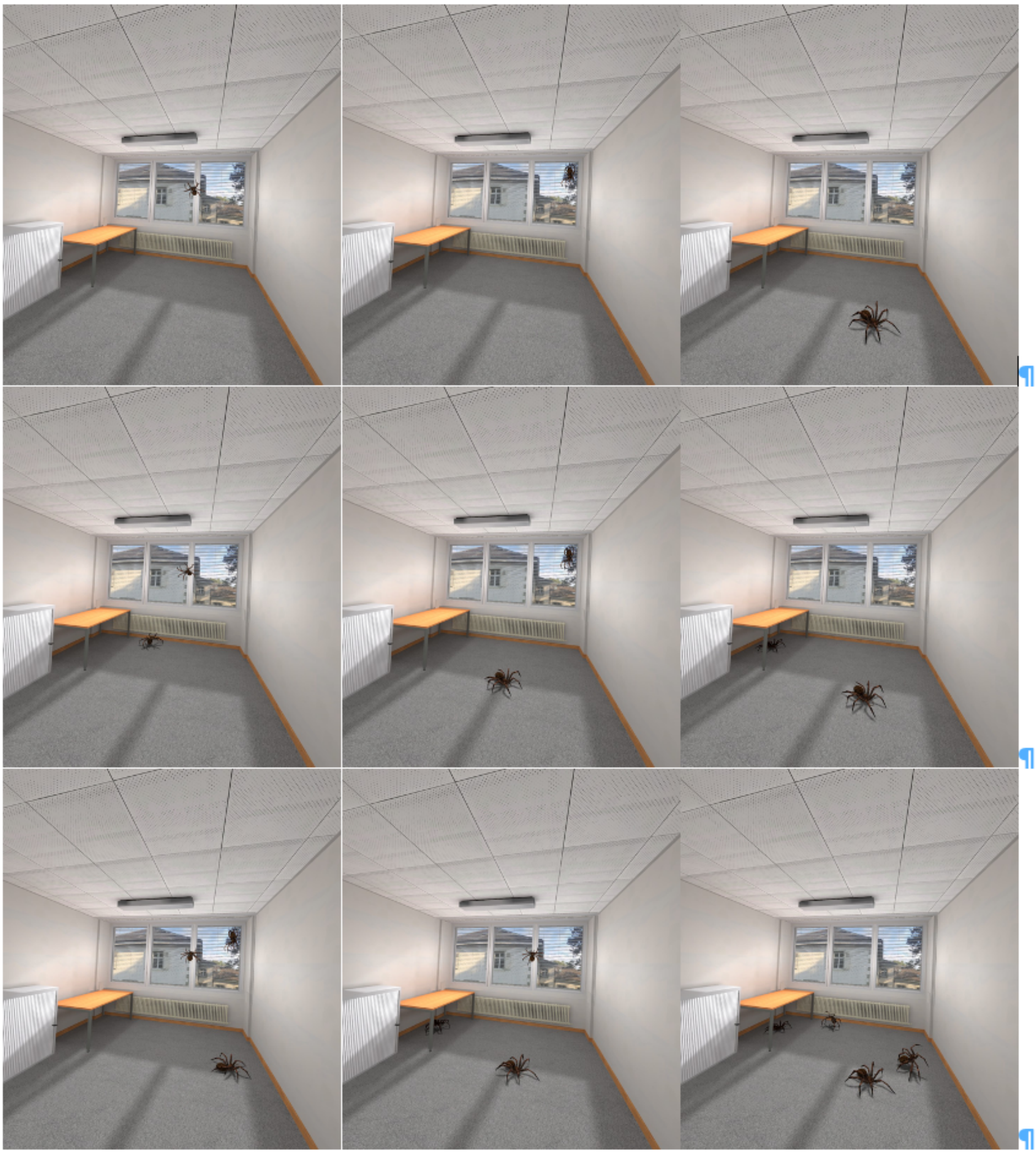

**Supplementary Figure SF2. Scenes 1–10 from the exposure therapy in VR.** The 10 scenes that were presented in a consecutive manner during exposure therapy in VR for 30 s per scene are displayed from left to right, starting with scene 1 and ending with scene 9 in the third row. Scene 10 corresponds to scene 1 to measure within-session fear reduction. During exposure therapy, the SUDS, HR, and SCL were measured in each of the 10 scenes at the beginning and end of each scene to quantify within-scene fear reduction.

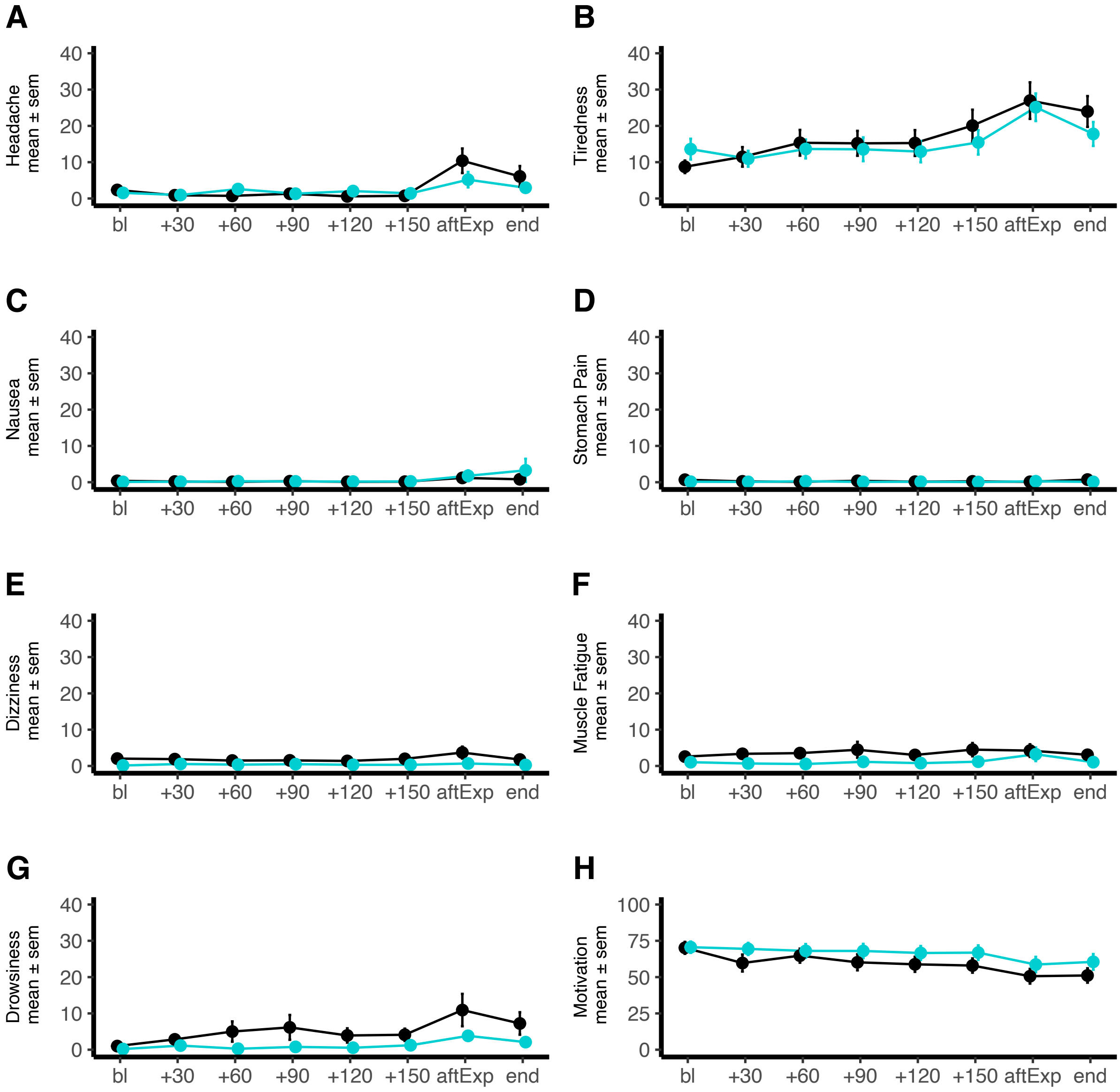


**Supplementary Figure SF3. Possible adverse events on vist 2 assessed by visual analogue scales (VAS).** The *x* axis represents the eight assessment timepoints, while the *y* axis depicts VAS ratings from 0 (not at all) to 100 (very strong). The black line represents the placebo + retrieval group, and the turquoise line represents the valproic acid + retrieval group. Displayed are means and standard errors of the mean. The *x* axis corresponds to the following time points baseline (bl), +30, +60, +90, +120, +150 min after medication intake, after exposure (aftExp), end of visit 2.

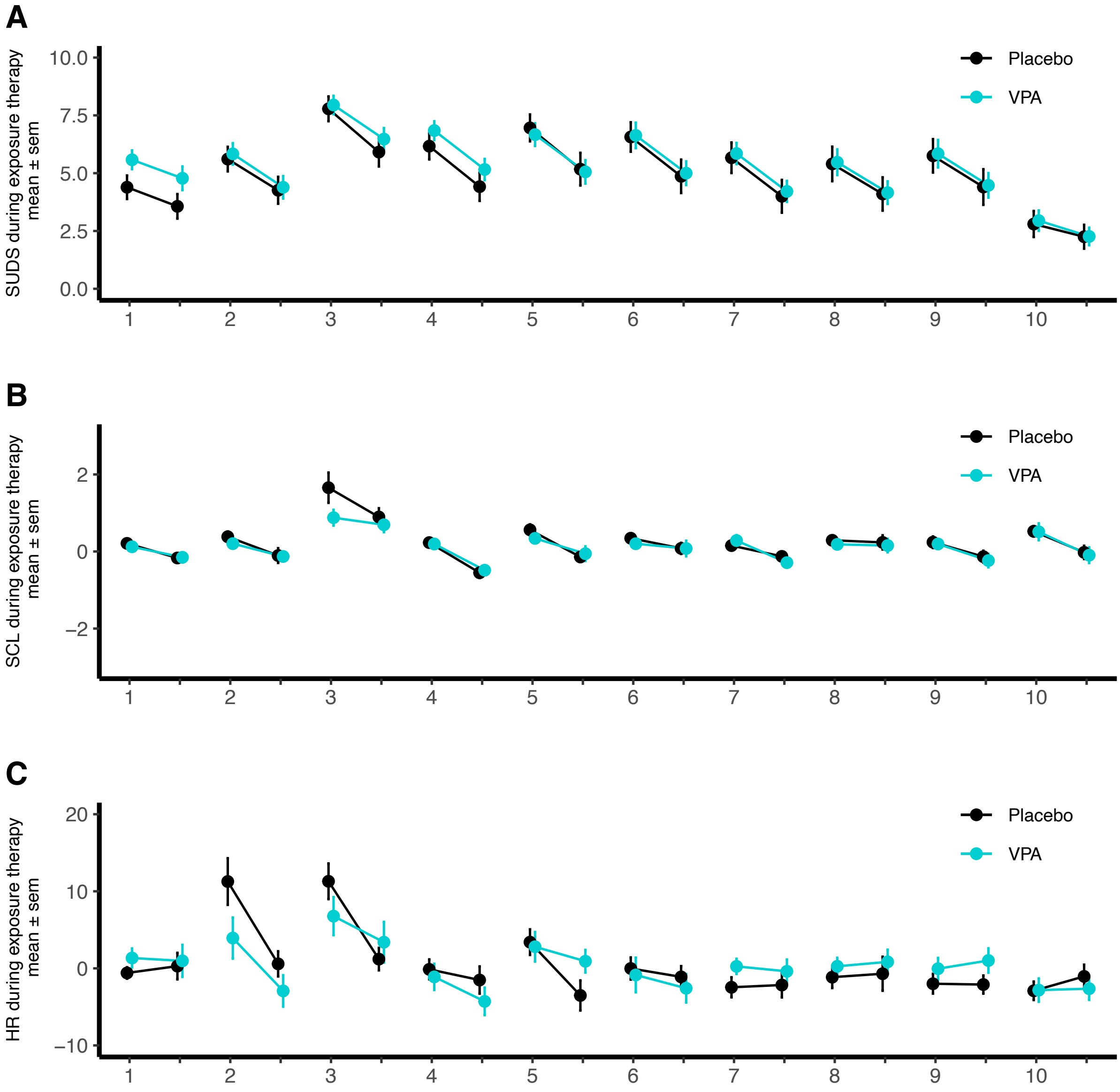


**Supplementary Figure SF4. Subjective Units of Distress Scale (SUDS) (A), skin conductance level (SCL) (B), and heart rate (HR) (C) during exposure therapy in VR.** The *x* axis represents the 10 VR scenes with values at the beginning and end of each scene. Scene 1 and 10 are identical to depict within-session change in the SUDS, SCL, and HR. The black line represents the placebo + retrieval group, and the turquoise line represents the valproic acid + retrieval group. Displayed are means and 95% confidence intervals.

### References

Arntz, A., Lavy, E., Van Den Berg, G., Van Rijsoort, S., 1993. Negative beliefs of spider phobics: A psychometric evaluation of the spider phobia beliefs questionnaire. Advances in Behaviour Research and Therapy 15, 257–277. https://doi.org/10.1016/0146-6402(93)90012-Q

Beck, A.T., Steer, R.A., Brown, G., 2011. Beck Depression Inventory–II. https://doi.org/10.1037/t00742-000

Blechert, J., Peyk, P., Liedlgruber, M., Wilhelm, F.H., 2016. ANSLAB: Integrated multichannel peripheral biosignal processing in psychophysiological science. Behav Res 48, 1528–1545. https://doi.org/10.3758/s13428-015-0665-1

Blumenthal, T.D., Cuthbert, B.N., Filion, D.L., Hackley, S., Lipp, O.V., Van Boxtel, A., 2005. Committee report: Guidelines for human startle eyeblink electromyographic studies. Psychophysiology 42, 1–15. https://doi.org/10.1111/j.1469-8986.2005.00271.x

Borkovec, T.D., Nau, S.D., 1972. Credibility of analogue therapy rationales. Journal of Behavior Therapy and Experimental Psychiatry 3, 257–260. https://doi.org/10.1016/0005-7916(72)90045-6

Bradley, M. M., & Lang, P. J, 2007. he International Affective Picture System (IAPS) in the study of emotion and attention., in: J. A. Coan & J. J. B. Allen (Ed.), Handbook of Emotion Elicitation and Assessment. Oxford University Press, pp. 29–46.

Dan-Glauser, E.S., Scherer, K.R., 2011. The Geneva affective picture database (GAPED): a new 730-picture database focusing on valence and normative significance. Behav Res 43, 468–477. https://doi.org/10.3758/s13428-011-0064-1

Fowles, D.C., Christie, M.J., Edelberg, R., Grings, W.W., Lykken, D.T., Venables, P.H., 1981. Publication Recommendations for Electrodermal Measurements. Psychophysiology 18, 232–239. https://doi.org/10.1111/j.1469-8986.1981.tb03024.x

Kennedy, R.S., Lane, N.E., Berbaum, K.S., Lilienthal, M.G., 1993. Simulator Sickness Questionnaire: An Enhanced Method for Quantifying Simulator Sickness. The International Journal of Aviation Psychology 3, 203–220. https://doi.org/10.1207/s15327108ijap0303_3

Lass-Hennemann, J., Michael, T., 2014. Endogenous cortisol levels influence exposure therapy in spider phobia. Behaviour Research and Therapy 60, 39–45. https://doi.org/10.1016/j.brat.2014.06.009

Laux, L, Glanzmann, P, Schaffner, P, Spielberger, CD, 1981. State-Trait-Angstinventar (STAI). Beltz, Weinheim.

Leiner, D.J., 2019. SoSci Survey.

Loos, E., Schicktanz, N., Fastenrath, M., Coynel, D., Milnik, A., Fehlmann, B., Egli, T., Ehrler, M., Papassotiropoulos, A., De Quervain, D.J.-F., 2020. Reducing Amygdala Activity and Phobic Fear through Cognitive Top–Down Regulation. Journal of Cognitive Neuroscience 32, 1117–1129. https://doi.org/10.1162/jocn_a_01537

Lüdecke, D., Ben-Shachar, M., Patil, I., Waggoner, P., Makowski, D., 2021. performance: An R Package for Assessment, Comparison and Testing of Statistical Models. JOSS 6, 3139. https://doi.org/10.21105/joss.03139

Noguchi, K., Gel, Y.R., Brunner, E., Konietschke, F., 2012. **nparLD** : An *R* Software Package for the Nonparametric Analysis of Longitudinal Data in Factorial Experiments. J. Stat. Soft. 50. https://doi.org/10.18637/jss.v050.i12

Pössel, P., Hautzinger, M., 2003. Dysfunktionale Überzeugungen bei Spinnenangst. Zeitschrift für Klinische Psychologie und Psychotherapie 32, 24–30. https://doi.org/10.1026/0084-5345.32.1.24

Rinck, M., Bundschuh, S., Engler, S., Müller, A., Wissmann, J., Ellwart, T., Becker, E.S., 2002. Reliabilität und Validität dreier Instrumente zur Messung von Angst vor Spinnen. Diagnostica 48, 141–149. https://doi.org/10.1026//0012-1924.48.3.141

Schiller, D., Monfils, M.-H., Raio, C.M., Johnson, D.C., LeDoux, J.E., Phelps, E.A., 2010. Preventing the return of fear in humans using reconsolidation update mechanisms. Nature 463, 49–53. https://doi.org/10.1038/nature08637

Schneider, S., Margraf, J., 2011. DIPS: diagnostisches Interview bei psychischen Störungen ; für DSM-IV-TR ; Handbuch, Interviewleitfaden, Protokollbogen ; mit CD-ROM, 4., überarb. Aufl. ed. Springer, Berlin Heidelberg.

Schubert, T.W., 2003. The sense of presence in virtual environments:: A three-component scale measuring spatial presence, involvement, and realness. Zeitschrift für Medienpsychologie 15, 69–71. https://doi.org/10.1026//1617-6383.15.2.69
Shah, D.A., Madden, L.V., 2004. Nonparametric Analysis of Ordinal Data in Designed Factorial Experiments. Phytopathology® 94, 33–43. https://doi.org/10.1094/PHYTO.2004.94.1.33
Shiban, Y., Brütting, J., Pauli, P., Mühlberger, A., 2015. Fear reactivation prior to exposure therapy: Does it facilitate the effects of VR exposure in a randomized clinical sample? Journal of Behavior Therapy and Experimental Psychiatry 46, 133–140. https://doi.org/10.1016/j.jbtep.2014.09.009
Spielberger, Charles D., Gorsuch, R. L., Lushene, R., Vagg, P. R., Jacobs, G. A., 1970. Manual for the State-Trait Anxiety Inventory. Consulting Psychologists Press, Palo Alto, CA.
Steyer, R., Schwenkmezger, P., Notz, P., Eid, M., 2017. Mehrdimensionalen Befindlichkeitsfragebogen. https://doi.org/10.1037/t12446-000
Szymanski, J., O'Donohue, W., 1995. Fear of Spiders Questionnaire. Journal of Behavior Therapy and Experimental Psychiatry 26, 31–34. https://doi.org/10.1016/0005-7916(94)00072-T
Wolpe, J., 2018. Subjective Units of Distress Scale. https://doi.org/10.1037/t05183-000
